# Toward a Disease Module for ME/CFS: A Network-Based Gene Prioritization

**DOI:** 10.1101/2025.04.13.25325733

**Authors:** Paolo Maccallini

## Abstract

**Background:** Myalgic Encephalomyelitis/Chronic Fatigue Syndrome (ME/CFS) is a debilitating condition with unclear etiology and no FDA-approved treatment. Recent studies suggest a possible genetic contribution to its pathogenesis.

**Objective:** This study aims to identify candidate genes for ME/CFS using both empirical evidence from genome-wide and next-generation sequencing studies on monogenic cases and computational expansion based on protein-protein interaction networks.

**Methods:** Twenty-two genes associated with ME/CFS were identified from relevant literature, including both common and rare variants. These genes were used as seeds in the STRING database to retrieve high-confidence interacting genes. A Random Walk with Restart (RWR) algorithm ranked 1063 candidate genes by their similarity to the seeds. The top 250 ranking genes were selected to define a disease module termed the ME/CFS module. This module was analysed for enrichment in metabolic pathways and disease associations.

**Results:** Enrichment analysis identified significant overlaps with sphingolipid metabolism and signaling, and energy-related pathways. Heme degradation, TP53-regulated metabolic genes, and thermogenesis were also identified as possibly contributing to the pathogenesis of ME/CFS. Overlaps with metabolic and neurodegenerative diseases were observed.

**Conclusion:** The ME/CFS module captures biologically plausible mechanisms underlying ME/CFS, with a particular focus on lipid and energy metabolism. It also provides a tool for filtering exome and genome data for the study of Mendelian cases of ME/CFS.

## 1 Introduction

Myalgic encephalomyelitis/chronic fatigue syndrome (ME/CFS) is a debilitating disorder of unknown cause, currently without FDA-approved treatments. It is defined by persistent fatigue, post-exertional symptom exacerbation, cognitive impairment, and orthostatic intolerance, lasting at least six months (1, 2). Although the pathophysiology remains unclear (3) multiple studies have identified abnormalities in blood metabolites (4–7), brain metabolism (8–10), aerobic capacity (11, 12), and evidence of central and systemic inflammation (10, 13–15). ME/CFS affects approximately 0.9% of the population (16) and most commonly begins in females under the age of forty (17). While the severity of symptoms fluctuates, remission is rare (1) and most patients do not return to their previous occupational status (18).

A study found significant excess ME/CFS relative risk among first to third-degree relatives of ME/CFS cases, suggesting a heritable contribution to the disease (19), in agreement with a previous work that reported an increased prevalence of ME/CFS among first-degree relatives of cases (20). Another study documented a significant rise in the prevalence of the disease in first to third-degree relatives of patients as well as in spouses/partners of cases, suggesting an interplay between genetic and environmental predisposing factors (21).

ME/CFS is among the phenotypes reported by participants of the UK Biobank database, a large-scale biomedical repository that includes genetic data on 500,000 UK citizens (22). An analysis of metadata generated from it showed a statistically significant association between 1208 females with self-reported ME/CFS and a regulatory region of chromosome 13 encompassing *SLC25A15*, the gene encoding ornithine transporter type I. The same analysis also indicated a possible association between the total cohort of 1659 ME/CFS patients (including 451 males) and an uncommon variant of *EBF3*, a transcription factor involved in B-cell differentiation, bone development, and neurogenesis (23, 24) (Table 1).

**Table 1.** Significant genetic associations for ME/CFS (UKBB.CFS) and chronic fatigue (UKBB.R53) from the analysis of a summary statistics generated from the UKBiobank (24). These associations were identified using standard GWAS analysis. In particular, a cut-off for significance for marginal regression of *p* < 5 × 10^−8^ was adopted for common variants (MAF above 0.05), while a more stringent cut-off of *p* < 1 × 10^−8^ was required for uncommon variants (MAF between 0.01 and 0.05). All the elements of this table, with the exclusion of the four pseudogenes, were used as seeds, along with the items of Table 2, for the generation of the ME/CFS module. An electronic version of this data is included in Supplementary Table S8.

| Primary gene list | Genes | Cases | Sex | Phenotype | Description | Ref. |
| --- | --- | --- | --- | --- | --- | --- |
| UKBB.CFS | *CYCSP34, SLC25A15, SUGT1P3, TPTE2P5, MRPS31* | 1208 | F | self-reported CFS (PHESANT) | Expression affected (eQTL) by two common variants (rs1536807, rs2017696) associated with phenotype in standard GWAS-type analysis ($p < 5.0E-08$). | (24) |
| UKBB.CFS | *CYCS, SUGT1, TPTE2* | 1208 | F | self-reported CFS (PHESANT) | One common variant affecting the expression of a corresponding coexpressed pseudogene is associated with self-reported ME/CFS. | - |
| UKBB.CFS | *EBF3* | 1659 | F/M | self-reported CFS (PHESANT) | Harbouring a rare variant (rs148723539) of possible regulatory function (score 5 on RegulomeDB) associated with phenotype in standard GWAS-type analysis ($p < 1.0E-08$). | (24) |
| UKBB.R53 | *ATP8B1* | 362 | M | ICD10: R53 | Expression affected (eQTL) by a common variant (rs62092652) associated with phenotype in standard GWAS-type analysis ($p < 5.0E-08$). | (24) |
| UKBB. R53 | *NUPR2, CHCHD2, SEPTIN14P24* | 456 | F | ICD10: R53 | Expression affected (eQTL) by an uncommon variant (rs182581502) associated with phenotype in standard GWAS-type analysis ($p < 1.0E-08$). | (24) |

Whole-genome sequencing (WGS) on 20 ME/CFS patients in the severe end of the spectrum reported an increased frequency of likely pathogenic and pathogenic variants (according to ACMG classification) in nine genes, most of which have been implicated in cognitive symptoms and neurological function (25) (Table 1).

Three cases of possible Mendelian ME/CFS have been reported. A putative causal variant was identified in a missense mutation on gene *NOS3* (c.3178G>A, p.E1060K), detected by whole-exome sequencing (WES) in a young woman with abnormal maximum effort study (low oxygen extraction, reduced cardiac output, low right-atrial pressure). This gene is associated with exercise intolerance in mice and is involved in vascular tone regulation (26). A structural variant spanning the AKR1C locus and detected by long-reads whole-genome sequencing (WGS) was proposed as causal in the case of a woman who experienced progressive mental and physical fatigue since the age of sixteen, after a viral infection. According to the authors, this variant disrupts the expression of *AKR1C1* (reduced) and *AKR1C2* (increased), leading to increased synthesis of inhibitory GABAergic neurosteroids and impaired Progesterone metabolism (27). A rare homozygous mutation on *SMPD1*, a gene associated with sphingomyelin lipidosis, was suggested as responsible for a significant increase in intracellular giant lipid droplet-like organelles in PBMCs in the case of a subject with extremely severe ME/CFS (28). The genes involved in these three cases are collected in Table 2.

**Table 2.** Causal or contributing genes identified in three cases of possible monogenic ME/CFS and twenty severely ill ME/CFS patients. The thirteen genes of this table were added to the nine genes of Table 1 (pseudogenes were excluded) and used as seeds for the generation of the ME/CFS module. An electronic version of this data is included in Supplementary Table S8.

| Primary gene list | Genes | Cases | Sex | Phenotype | Description | Ref. |
| --- | --- | --- | --- | --- | --- | --- |
| MMECFs | *NOS3* | 1 | F | Chronic exercise intolerance and orthostatic hypotension | Harboring a heterozygous missense mutation (c.3178G>A - p.Glu1060Lys - detected by WES), proposed as causal in this case. | (26) |
| MMECFs | *AKR1C1, AKR1C2* | 1 | F | ME/CFS with atypical response to steroid medications | Impacted by a large structural variant detected with WGS long read sequencing, proposed as causal in this case. | (27) |
| MMECFs | *SMPD1* | 1 | M | Very severe ME/CFS | Housing a homozygous missense mutation (c.808G>A - p.Gly270Ser) detected by WES, proposed as contributing to this case. | (28) |
| SIPS | *ACADL, BRCA1, CFTR, COX10, HABP2, MFRP, PCLO, PRKN, ZFPM2* | 20 | F/M | ME/CFS | Increased frequency of pathogenic and likely-pathogenic variants (ACGM classification) in severe ME/CFS patients. | (25) |

Given the limited genetic research on ME/CFS, it is reasonable to expand existing knowledge by leveraging protein–protein interaction (PPI) networks. In this study, we present a gene list derived from published data on both Mendelian and idiopathic ME/CFS. These seed genes were expanded using STRING, a PPI database that integrates evidence from gene expression, proteomics, curated databases, and text mining (29).

The method presented here is grounded in the literature briefly discussed below. Genes involved in the same disease or phenotype show a significant tendency to interact with one another or, in other words, to be neighbours in the corresponding protein interaction network (30–32). This property defines phenotype-specific subgraphs known as disease modules, where unknown members can be inferred from a few known genes, or seeds (33). Importantly for the present study, it has been discovered that mutations associated with rare monogenetic diseases identify pathways involved in the pathogenesis of common diseases with matched phenotypes (34). A recent large-scale analysis confirmed that genes disrupted in Mendelian disorders frequently harbor common variants associated with related complex traits (35). This supports the integration of both GWAS and next-generation sequencing (NGS) data to build biologically informed seed sets for module discovery.

While genes associated with the same disease tend to be clustered, there may be no direct edge between them. This implies that not all interactors with seeds are relevant. Several mathematical approaches of network analysis have been proposed to prioritize the candidate genes, and random walk with restart (RWR) has shown to be effective in addressing this problem (36, 37).

In this study, we retrieve seed genes from the literature on both idiopathic ME/CFS and Mendelian cases compatible with ME/CFS diagnostic criteria. These were expanded using STRING (interaction score ≥ 0.7) and then ranked using RWR. The resulting list, referred to as the ME/CFS module, was analyzed for functional enrichment and is proposed as a tool for prioritizing candidate genes in suspected Mendelian ME/CFS.

## 2 Methods

### 2.1 Analysis of the UK Biobank dataset

Some of the phenotypes included in the UK Biobank database are collected in three files (one for each sex group) that can be found on the web page of the Neale Lab (http://www.nealelab.is/uk-biobank). We searched with the keywords: fatigue, myalgic, encephalomyelitis, fibromyalgia, post, post-viral, POTS, tachycardia, dysautonomia, dysautonomic, and autonomic. This query selected the phenotypes collected in Table 4. We downloaded the corresponding summary statistics from the Neale Lab webpage.

We searched for statistically significant associations between variants and phenotypes according to the following selection criteria. Variants with a minor allele frequency (MAF) below 0.001 or an expected number of minor alleles in cases below 25 (low confidence status) were removed. Filtering by Hardy-Weinberg equilibrium (HWE) was performed by inspecting column 16 of `variants.tsv`. Variants with HWE p-values < 10^−6^ in the total sample were considered potential genotyping errors (38). In the study of the associations between variants and phenotypes, a significant threshold of 5 × 10^−8^ was used for variants with MAF above 0.05 (common variants), while a more stringent threshold of 1 × 10^−8^ was applied for uncommon variants (MAF between 0.01 and 0.05) (39).

For example, the filtering process for females with self-reported ME/CFS generated the set of eleven common variants in Table S 1. When there was more than one significant association for a given phenotype (as in the above example), we proceeded with fine mapping. To do so, we need an estimate of linkage disequilibrium (LD) between the candidate variants. We interrogated LDlink (40) and called function `LDhap()`of the R package *LDlinkR* (41) with reference genome GRCh37, GBR reference population, and the list of rsids from the previous step (second column of Table S 1, in the example). The output for variants of Table S 1 is the one reported in Table S 2. We then calculated the LD matrix, with each element at position *i*, *j* defined as:

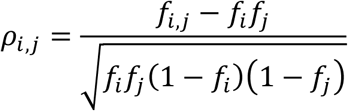

where *f*_*i*_ is the frequency of the alternative allele of the control group at position *i*, *f*_*j*_ has the same meaning for position *j*, and *f*_*i*,*j*_is the frequency of the haplotype defined by the presence of the alternative allele at positions *i* and *j*, in the reference population (42). Value *ρ*_*i*,*j*_ is a measure of LD between loci *i*, *j* (43). The correlation matrix for the eleven variants of the example is reported in Table S 3. Next, we input the LD matrix in function `fitted_rss()`of the R package *SusieR*, along with the Z scores of the variants, derived from the regression coefficients and standard errors. *SusieR* computes posterior inclusion probability (PIP) values: *PIP*_*i*_ represents the conditional probability that the regression coefficient of the joint model relative to variant *i* is different from zero, given the data (42). We also interrogated RegulomeDB to identify regulatory variants among those associated with traits and their corresponding target genes. RegulomeDB integrates data from GWAS results on noncoding regions, regulatory regions classified by the ENCODE Project, manually curated loci known to be involved in regulatory functions, and loci that explain variation in the expression level of mRNA, known as Expression Quantitative Trait Loci (eQTLs) (44).

This procedure suggests an association of the phenotypes of interest with four pseudogenes. Although traditionally considered non-functional, pseudogenes can be transcribed into mRNA and, in some cases, translated into non-canonical proteins (45). Coexpression between pseudogenes and their corresponding parental genes has been documented, supporting the hypothesis that pseudogenes may exert regulatory effects on their parental counterparts (46). Pseudogenes have been implicated in several human diseases, and a variety of mechanisms have been described, including interference with parental gene expression (47).

### 2.2 Study of Pseudogenes

Pseudogenes are not included in the STRING database; therefore, we used their corresponding parental genes as a proxy, provided that significant coexpression was observed between each pseudogene and its parental gene. Parental gene assignments were based on the NCBI Gene database. To test coexpression, we used GTEx release V10 (downloaded from the GTEx Portal, https://gtexportal.org, accessed on 2025-03-29), which includes TPM readings for each sample. We also downloaded the corresponding annotation with samples classified by tissue type. For each pseudogene-parental gene pair, we calculated the Pearson correlation coefficient for expression values within each tissue, along with the associated *p*-value. A conservative Bonferroni correction was applied to control for multiple testing: we used a significance threshold *p* < 4.1 ⋅ 10^−13^, calculated as *α*⁄*m*, with *α* = 0.05 and *m* given by the number of possible gene pairs within the GTEx database (∼1.74 ⋅ 10^9^) multiplied by 70 (the number of tissues available). Only tissues with more than twenty samples were included in the study of coexpression. It is important to note that coexpression is a necessary but not a sufficient condition for interaction between a pseudogene and its parental gene.

### 2.3 List expansion and Gene Graph

Genes listed in Table 1 and Table 2 were used as seeds for interrogating the STRING database after substituting pseudogenes with their corresponding parental genes. A minimum probability of interaction of 0.7 – labeled as *high confidence* on STRING’s website – was required. For each seed, we collected all the interacting genes, hereafter referred to as candidate genes, along with the associated STRING interaction scores. We used version 12.0 of the STRING database, downloaded locally.

After expansion, the four gene lists containing the seeds – referred to as Primary Gene Lists (PGLs) and collected in Table 1 and Table 2 – are transformed into their respective Extended Gene Lists (EGLs). This procedure yielded a symmetric square matrix, where the element at row *i* and column *j* is the STRING interaction score between gene *i* and gene *j.* This matrix can be interpreted as the adjacency matrix of a weighted, undirected graph. The union of all EGLs is termed the merged gene list (MGL). Refer to Table 3 for a summary of terminology and definitions.

**Table 3.** Terminology and definitions adopted in the present study.

| Name | Description |
| --- | --- |
| Primary gene list | Each unique item in the first column of Table 1 and Table 2 is a primary gene list (PGL). It includes genes directly associated with the phenotype ME/CFS in a cohort of patients (Table 1) or studies of a single individual (Table 2). |
| Expanded gene list | A PGL after expansion via the STRING database becomes an expanded gene list (EGL). An EGL includes both the original seeds and genes that show a significant interaction with them, according to the STRING database. |
| Merged gene list | The union of all the available EGLs is called the merged gene list (MGL). |
| Seed gene | Each gene of a PGL is referred to as a seed gene. A seed gene can belong to more than one PGL. In this study, a total of twenty-two seed genes were included. |
| Candidate gene | Each gene with significant protein-protein interaction with a seed gene is referred to as a candidate gene. Note that a seed gene can also be a candidate gene and that a candidate gene can belong to more than one EGL. |
| ME/CFS module | Ranking and filtering of the MGL by random walk with restart gives rise to a smaller list of genes, called the ME/CFS module, which is then used for over-representation analysis and is proposed as a filtering tool for the study of exomes and genomes of probands with suspected Mendelian ME/CFS. |

**Table 4.**
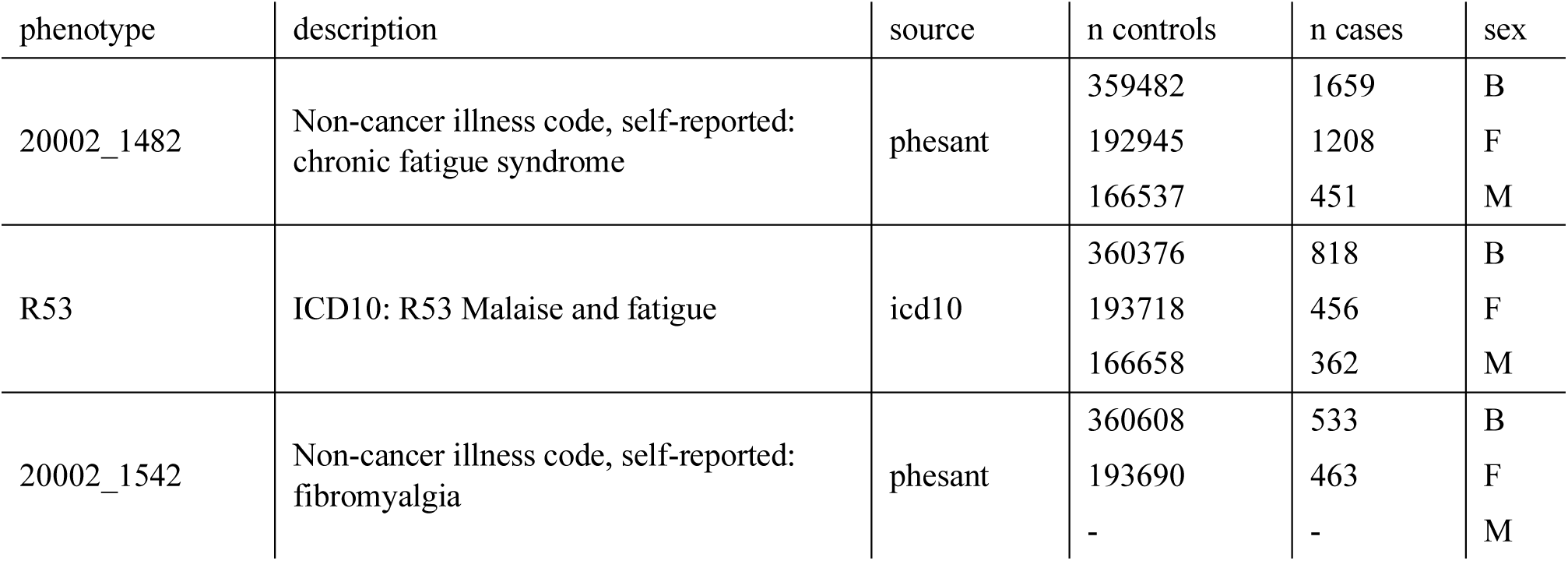
Phenotypes considered in the UK Biobank. B: both sexes; F: females; M: males. Phesant: phenotypes that are automatically processed using a modified version of the software PHESANT (90). For each phenotype, the number of females (F) and males (M) are indicated, along with the total number of individuals (B, for both sexes).

| phenotype | description | source | n controls | n cases | sex |
| --- | --- | --- | --- | --- | --- |
| 20002_1482 | Non-cancer illness code, self-reported: chronic fatigue syndrome | phesant | 359482 | 1659 | B |
|  |  |  | 192945 | 1208 | F |
|  |  |  | 166537 | 451 | M |
| R53 | ICD10: R53 Malaise and fatigue | icd10 | 360376 | 818 | B |
|  |  |  | 193718 | 456 | F |
|  |  |  | 166658 | 362 | M |
| 20002_1542 | Non-cancer illness code, self-reported: fibromyalgia | phesant | 360608 | 533 | B |
|  |  |  | 193690 | 463 | F |
|  |  |  | - | - | M |

To construct and visualize the graph from the adjacency matrix, we used the R package *igraph* (48). This graph is referred to as the Gene Graph (GG) to distinguish it from the List Graph (LG), described below.

### 2.4 List Graph

Each gene of the MGL – whether a seed or a candidate – can belong to more than one EGL. This overlap creates connections between different EGLs, which can be captured in a symmetric square matrix, where each element represents the number of genes shared between a given pair of EGLs. This matrix can be interpreted as the adjacency matrix of a weighted undirected graph, referred to as the List Graph (LG).

### 2.5 Gene scoring algorithm by a random walk with restart

As mentioned, the MGL was used to build a weighted undirected graph with one node for each gene and one edge for each pair of genes with a STRING score greater than 0.7. The edge weight corresponds to the STRING score.

To compute a global score of interaction between candidate genes and seeds, we applied random walk with restart (RWR) as described in (36), by custom R script. Briefly, given a connected weighted undirected graph *G*(*V*, *E*), with a set of vertices *V* = {*v*_1_, *v*_2_, …, *v*_*N*_} and a set of edges *E* = {(*v*_*i*_, *v*_*j*_)|*v*_*i*_, *v*_*j*_ ∈ *V*}, where *a*_*ij*_ is the weight of edge (*v*_*i*_, *v*_*j*_), let *A* be the adjacency matrix associated with *G*. This matrix is symmetric, with diagonal elements equal to zero. The RWR is then defined by the stochastic process:

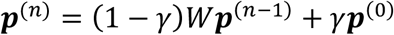

where *γ* is the restart probability, ***p***^(*n*)^is the probability distribution at iteration *n*, and *W*is the column-normalized form of *A*. It can be shown that ***p***^(*n*)^converges to the stationary distribution:

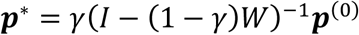

The standard approach is to initialize ***p***^(0)^ such that each seed gene has equal probability, while all candidate genes have a starting probability of zero. In our case, a starting probability of 1/21 was assigned to each seed gene except for *AKR1C1* and *AKR1C2.* They were each initialized to 0.5/21 to avoid overrepresentation due to their proximity in the STRING network.

The asymptotic probability ***p***^∗^, which represents a measure of the connectivity across the network of each node with the seeds, was used to rank all candidate genes in the MGL. We set the restart probability to *γ* = 0.7, following prior recomendations (49). Next, we scaled the scores between 0 and 1. The top 250 ranked candidates were selected and merged with the 22 original seeds to define the ME/CFS module.

### 2.6 Enrichment analysis

To extract biological meaning from the ME/CFS module, we performed over-representation analysis (ORA) using several curated databases.

Enrichment against the Kyoto Encyclopedia of Genes and Genomes (KEGG) was performed using the `enrichKEGG()`function of package *ClusterProfiler* (50, 51). We also queried the Reactome pathway database – an expert-curated resource of biological pathways – using the `enrichPathway()`function from the *ReactomePA* package (52, 53). To identify disease associations, we used the `enrichDO()`function from the *DOSE* package (54), which is based on the Disease Ontology (DO) database (55). Finally, to assess tissue-specific expression, we applied the `teEnrichment()`function from the *TissueEnrich* package (56).

In all enrichment analyses, *p*-values were adjusted using the Benjamini-Hochberg (BH) procedure to control for false discovery rate (57). These methods are summarised in Table 7.

### 2.7 Randomly generated genetic lists

To test whether the ME/CFS module reflects a biologically meaningful structure rather than a random assembly of genes, we generated random sets of human genes with the same structure as the set described in Table 1 and Table 2. Each random set includes the same number of seeds for each PGL. Expansion by STRING was performed as described. A total of 600 random MGLs were generated. For each random MGL, we computed topological features of both the GG and the LG, including the number of nodes and connected components for GG; the number of edges, total weight, mean degree, and number of connected components for LG.

To evaluate whether the MGL obtained for ME/CFS deviated significantly from the randomly generated MGLs, we applied three strategies. One-tailed non-parametric tests were performed to compare each topological metric of the ME/CFS MGL against the corresponding empirical distribution for the 600 random MGLs. Next, we performed principal component analysis (PCA) on the sample of random lists plus the ME/CFS list and verified whether the ME/CFS MGL falls outside the 95% confidence ellipse in the PC1-PC2, PC1-PC3, PC2-PC3 planes. We also tested if the ME/CFS list is included among the 5% outliers of the set of MGLs. To do so, we employed the local outlier factor (LOF) algorithm (58). We set 30 as the minimum value for minPts (it represents the minimum size of a cluster for the determination of local outliers), and we chose 50 as the maximum value, considering that it represents the size of the larger cluster that can be regarded as a cluster of outliers. For each value of minPts from 30 to 50, we calculated the maximum LOF of each MGL (including the ME/CFS one) and then selected as outliers those elements that reach the top 5% LOF values. The calculation of LOF was performed by function `lof()`of package *dbscan*.

### 2.8 Genetic Annotation

We annotated each gene of the ME/CFS module with its probabilities of haploinsufficiency (pHaplo) and triplosensitivity (pTriplo) (59). Probabilities were downloaded from Zenodo (60), at record 6347673. We also included the DOMINO score, a probabilistic estimate of dominant inheritance (pDI) (61). We compared the distributions of pDI, pHaplo, and pTriplo between the ME/CFS module genes and the human genome. The comparison was conducted using two-sided Wilcoxon rank-sum tests.

## 3 Results

This pipeline began by identifying genes associated with ME/CFS and ICD-10 code R53 (Chronic Fatigue) using data from the UK Biobank. This analysis yielded nine genes, including three pseudogenes – *CYCSP34, SUGT1P3,* and *TPTE2P5* – whose expression was found to correlate significantly with their respective parental genes across multiple tissues (Table 6, Figure S 2), suggesting a potential regulatory interaction. Since pseudogenes are not represented in the STRING database, they were replaced by the corresponding parental genes for all subsequent analyses. The nine protein-coding genes identified through this prioritization process are listed in Table 8, along with a brief description of their biological functions and known associations with disease.

**Table 5.** Probability of dominant pattern of inheritance (pDI), of haploinsufficiency (pHaplo), and triplosensitivity (pTriplo) among the genes of the ME/CFS module and of the human genome. Data are indicated as medians with standard deviations in brackets. Probabilities of dosage sensitivity, pHaplo and pTriplo, are developed by the Talkowski lab (59) and predict whether a gene is haploinsufficient or triplosensitive, respectively. Analogously, pDI is a probability of dominant inheritance (61). A double-tailed Wilcoxon test was performed to compare the median probabilities among the genes of the ME/CFS module and the corresponding medians among the totality of human genes.

| variable | ME/CFS module | Human genes | p-value |
| --- | --- | --- | --- |
| pDI | 0.18 (0.37) | 0.16 (0.31) | 3.20E-02 |
| pHaplo | 0.57 (0.29) | 0.4 (0.3) | 2.89E-07 |
| pTriplo | 0.48 (0.29) | 0.37 (0.29) | 1.42E-04 |

**Table 6.** Correlations between gene expression of pseudogene-parental gene pairs by tissue. Gene expression data for seventy tissues, expressed as TPM, were retrieved from release V10 of the GTEx database. Pearson correlation coefficients and corresponding *p*-values were calculated by function `cor.test()`(R programming language). We considered as significant *p*-values below 0.05⁄*m*, with *m* given by the number of gene pairs within the GTEx Database multiplied by the total number of available tissues. The number of genes in GTEx is *n* = 59033. Therefore, the number of pairs is 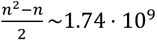 and *m*∼122 ⋅ 10^9^. The cut-off for significance is then *p* < 4.1 ⋅ 10^−13^.

| pair | Tissue | Pearson | samples | p-value |
| --- | --- | --- | --- | --- |
| TPTE2-TPTE2P5 | Brain - Hypothalamus | 0.56 | 257 | 2.17E-22 |
|  | Brain - Anterior cingulate cortex (BA24) | 0.48 | 233 | 8.67E-15 |
|  | Brain - Nucleus accumbens (basal ganglia) | 0.47 | 285 | 9.66E-17 |
|  | Brain - Caudate (basal ganglia) | 0.41 | 300 | 6.45E-14 |
|  | Pancreas | 0.40 | 362 | 2.21E-15 |
|  | Cells - Cultured fibroblasts | 0.37 | 652 | 2.78E-22 |
|  | Heart - Atrial Appendage | 0.36 | 461 | 3.21E-15 |
|  | Breast - Mammary Tissue | 0.35 | 514 | 1.16E-16 |
|  | Muscle - Skeletal | 0.34 | 818 | 9.46E-24 |
|  | Nerve - Tibial | 0.32 | 670 | 1.50E-17 |
|  | Whole Blood | 0.30 | 803 | 9.19E-18 |
| CYCS-CYCSP34 | Whole Blood | 0.53 | 803 | 3.59E-58 |
|  | Brain - Caudate (basal ganglia) | 0.47 | 300 | 3.23E-18 |
|  | Brain - Nucleus accumbens (basal ganglia) | 0.46 | 285 | 2.23E-16 |
|  | Cells - Cultured fibroblasts | -0.38 | 652 | 2.57E-23 |
| SUGT1-SUGT1P3 | Brain - Nucleus accumbens (basal ganglia) | 0.77 | 285 | 1.02E-56 |
|  | Brain - Caudate (basal ganglia) | 0.70 | 300 | 1.09E-44 |
|  | Kidney - Cortex | 0.67 | 104 | 5.68E-15 |
|  | Brain - Putamen (basal ganglia) | 0.66 | 254 | 9.42E-34 |
|  | Brain - Hypothalamus | 0.55 | 257 | 1.90E-21 |
|  | Brain - Anterior cingulate cortex (BA24) | 0.54 | 233 | 8.60E-19 |
|  | Brain - Hippocampus | 0.53 | 255 | 3.98E-20 |
| SUGT1-SUGT1P3 | Pancreas | 0.52 | 362 | 3.17E-26 |
|  | Brain - Cerebellum | 0.51 | 266 | 9.75E-19 |
|  | Brain - Frontal Cortex (BA9) | 0.50 | 269 | 1.79E-18 |
|  | Brain - Cortex | 0.49 | 270 | 1.27E-17 |
|  | Breast - Mammary Tissue | 0.47 | 514 | 1.03E-29 |
|  | Heart - Left Ventricle | 0.41 | 452 | 7.36E-20 |
|  | Whole Blood | 0.36 | 803 | 1.72E-25 |
|  | Muscle - Skeletal | 0.27 | 818 | 4.87E-15 |
| SEPTIN14-SEPTIN14P24 | Prostate | 0.49 | 282 | 2.18E-18 |

**Table 7.** Methods for over-representation analysis. This table indicates the R functions employed for over-representation analysis and the corresponding packages. Also, the databases used by each function are indicated, with a brief description of their type of terms. The citations in the last column refer to the packages. The references for the databases are as follows: KEGG (50), Reactome (52), DO (55), Human Protein Atlas (91).

| Function | R Package | Database | Purpose | Ref |
| --- | --- | --- | --- | --- |
| `enrichKEGG()` | *ClusterProfile* | KEGG | Metabolic and signaling pathways | (51) |
| `enrichPA()` | *ReactomePA* | Reactome | Cell signaling, metabolism, disease pathways | (53) |
| `enrichDO()` | *DOSE* | Disease Ontology | Disease association | (54) |
| `teEnrichment()` | *TissueEnrich* | Human Protein Atlas | Tissues | (56) |

**Table 8.** Genes associated with Chronic Fatigue (ICD10:R53) and self-reported ME/CFS. Each gene was identified through UK Biobank analysis and used as seeds for the definition of the ME/CFS disease module. The table reports the phenotype used for association (ME/CFS or ICD10:R53), a brief description of the gene’s known function, and relevant literature.

| Gene | Phenotype | Description |
| --- | --- | --- |
| *ATP8B1* | ICD10:R53 | Encodes a translocase that transports phospholipids from one side of the cell membrane to another. This gene is deficient in patients with intrahepatic cholestasis, and *ATP8B1* knockdown in hepatocytes leads to an increase in oxidative phosphorylation (92). |
| *CHCHD2* | ICD10:R53 | It is a small protein localized in the mitochondrial intermembrane space. Mutations in this gene have been linked to both Mendelian and idiopathic Parkinson's disease (93). |
| *CYCS* | ME/CFS | Encodes cytochrome c, a key component of the mitochondrial electron transport chain. Mutations in this gene have been associated with a reduction in mitochondrial respiratory rate, increased apoptosis, and thrombocytopenia (94). |
| *EBF3* | ME/CFS | Encodes a transcription factor involved in B-cell differentiation, bone development, and neurogenesis. It localizes to the nucleus. Missense mutations on this gene cause intellectual disability and ataxia with a dominant autosomal pattern of inheritance (95). |
| *MRPS31* | ME/CFS | Encodes a mitochondrial ribosomal protein, part of the mitochondrial translation machinery (96). |
| *NUPR2* | ICD10:R53 | Involved in cellular response to starvation, it is regulated by *TP53* (97). |
| *SLC25A15* | ME/CFS | Encodes ornithine transporter type I, which transports ornithine across the inner mitochondrial membrane as part of the urea cycle. Mutations in this gene cause hyperornithinemia-hyperammonemia-homocitrullinuria syndrome, a condition characterised by lethargy and coordination problems where the ammonia degradation is impaired (98). |
| *SUGT1* | ME/CFS | Participates in kinetochore functions and interacts with heat shock protein 90 for the regulation of inflammasome activity (99). |
| *TPTE2* | ME/CFS | Encodes a membrane phospholipid phosphatase. It has been associated with total brain volume and fatty acid metabolism in GWAS studies (100, 101). |

This set was combined with nine genes carrying recurrent pathogenic or likely pathogenic variants – classified according to ACMG guidelines – in a cohort of 20 severely ill ME/CFS patients (25). Additionally, four genes previously proposed as causal in three independent cases of suspected Mendelian ME/CFS (26–28) were included, resulting in a final seed list of 22 genes divided into four PGLs (Table 1, Table 2).

For each one of the 22 genes (referred to as seeds), all interacting partners with a STRING confidence score above 0.7 (classified as high-confidence) were retrieved using a local installation of the STRING database, version 12.0 (29). This process yielded 1063 candidate genes. To complete the interaction network, all missing edges between the total 1085 genes were retrieved from STRING, resulting in a weighted, undirected graph of 20013 edges (Figure 1). The set of all nodes is referred to as the merged gene list (MGL).

**Figure 1.**
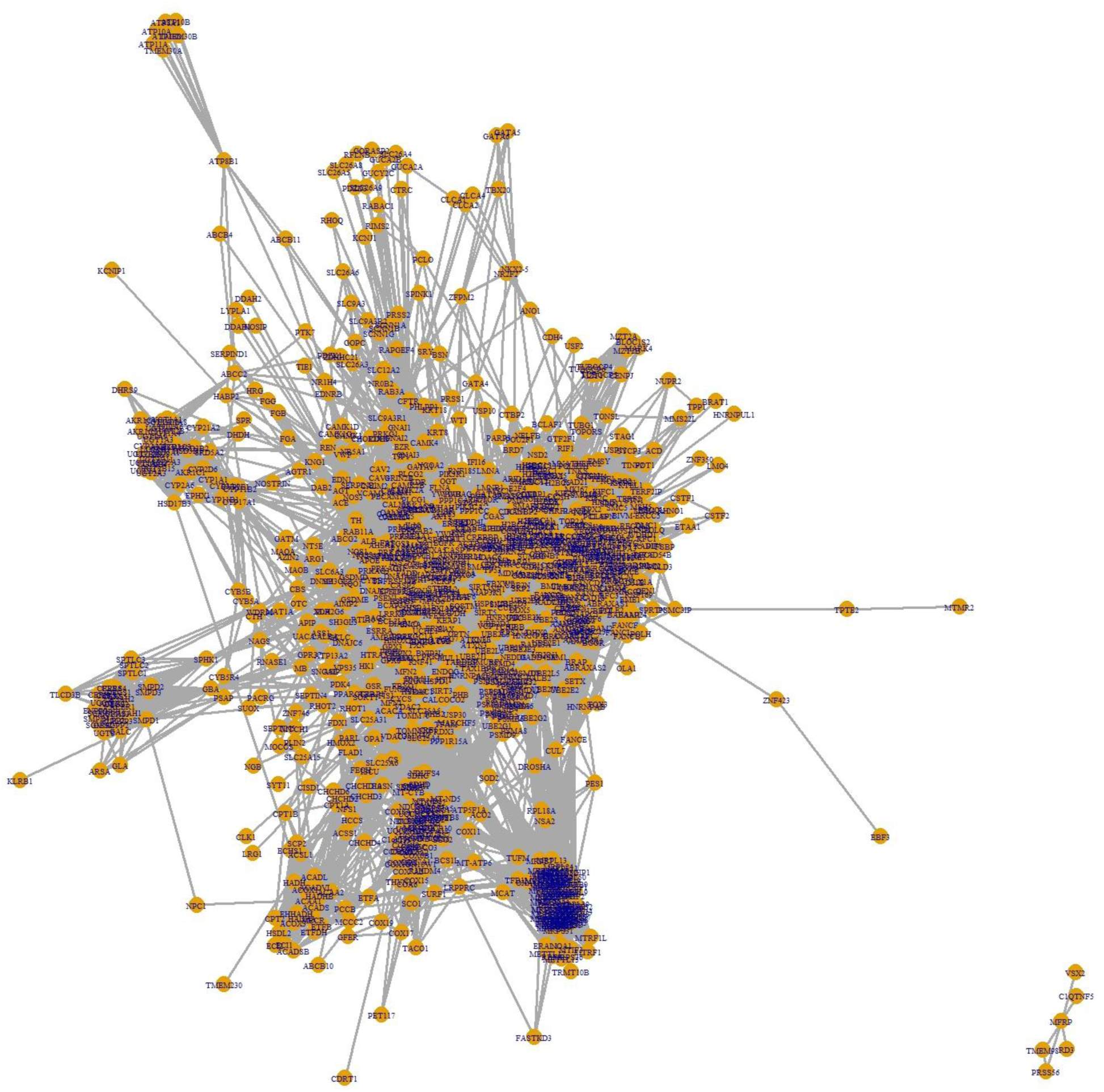
Gene Graph (GG). Weighted undirected graph associated with the merged gene list (MGL). There is a node for each of the 1085 genes (22 seed genes and 1063 candidates) and an edge for each pair of genes with a STRING probability of interaction greater than 0.7. This plot was generated using the R package *igraph* (48). All but six genes are included in a single connected component. On the bottom-right of the figure, the smallest component composed of seed MFRP (from the primary gene list SIPS) and its five interacting genes predicted by STRING.

Next, we applied the Random Walk with Restart (RWR) algorithm to the MGL to assign a connectivity score to each gene based on its proximity to the seeds in the network. Initial probabilities were evenly distributed among the seeds, with minor adjustments to account for redundancy (see Methods). The top 250 ranked candidate genes, together with the original 22 seeds, define the ME/CFS module, which was used for downstream enrichment analysis.

Several structural and topological features of the gene and list graph were calculated and compared against the same metrics computed for 600 randomly generated MGLs with matching characteristics. All computational analyses were carried out in R version 4.4.1. The detailed results of these steps are presented in what follows.

As part of our pipeline, we conducted a custom analysis of summary statistics from the UK Biobank to identify variants significantly associated with phenotypes listed in Table 4. In the analysis of variants with MAF above 0.05, we observed a significant association in females with self-reported ME/CFS involving a block in LD on chromosome 13, spanning positions 41,353,297 to 41,404,706 (hg19), which fully encompass the gene *SLC25A15*, which encodes ornithine transporter type I (Table S 1). Fine mapping analysis pinpointed rs11147812 (13:41404706:T:C) as the most likely causal variant within this region. This variant – which falls within pseudogene *TPTE2P5* – is associated with the whole sample (males and females) at 5 × 10^−8^ < *p* < 5 × 10^−7^. This same locus was independently identified by a Norwegian group, who reported a similar association at a higher *p*-value (9 × 10^−6^) using separately generated summary statistics from the UK Biobank (62)g.

Among the variants in Table S 1, we selected rs1536807 and rs2017696 based on their high probability of having regulatory effects. Genes significantly influenced by these variants, as supported by eQTL data from RegulomeDB, were manually retrieved, yielding the following: *CYCSP34, SLC25A15, MRPS31, SUGT1P3,* and *TPTE2P5*.

In males diagnosed with chronic asthenia not otherwise specified (ICD10: R53), we identified a significant association with a region in high LD on chromosome 18, spanning positions 55,452,281 to 55,460,845 (hg19). Within this segment, only one variant reached genome-wide significance (*p* < 5 × 10^−8^): rs62092652 (18:55454761:G>A). This variant was identified as the most likely causal locus based on PIP analysis across the region. Annotation from eQTL databases indicates that rs62092652 lies in a regulatory region of *ATP8B1*.

We extended our analysis to uncommon variants (MAF between 0.01 and 0.05), applying a more stringent genome-wide significance threshold (*p* < 1 × 10^−8^). This approach revealed a significant association between ME/CFS (both sexes combined) and rs148723539 (10:131677233:G>A, hg19), a putative regulatory variant within the gene *EBF3* (*p* = 2.2 × 10^−9^). Additionally, in females diagnosed with ICD10: R53, we identified rs182581502 (7:56492485:G>T, hg19) as significantly associated with the phenotype (*p* = 5.4 × 10^−9^). The alternative allele of this variant, located in the intronic region of pseudogene RP13-492C18.2, was more frequent in patients than in controls. eQTL analysis using RegulomeDB indicates that rs182581502 has a regulatory effect on the expression of NUPR2, SEPTIN14P24, and CHCHD2. No significant associations were found in the male-only subgroup.

No genome-wide significant variants were detected for the fibromyalgia phenotype.

The previous step suggested an association between female ME/CFS subjects and the pseudogenes *CYCSP34*, *SUGT1P3*, and *TPTE2P5*, and an additional association between females with phenotype R53 and the pseudogene *SEPTIN14P24*. We then assessed coexpression between each pseudogene and its respective parental gene, as shown in Table 6 (correlation coefficients by tissue) and Figure S 2 (expression by tissue). Notably, the pair *SEPTIN14*-*SEPTIN14P24* showed statistically significant coexpression only in prostate tissue; since this association was found in a female cohort, we excluded *SEPTIN14* from the seed list. In contrast, *CYCSP*, *SUGT1*, and *TPTE2* exhibited significant coexpression with their respective pseudogenes across multiple tissues and were, therefore, considered suitable proxies for the pseudogenes in downstream network analyses. They were included as seed genes in the primary gene list UKBB.CFS (Table 1).

Genes listed in Table 1 and Table 2 were used as seeds for interrogation of the STRING database, following the substitution of pseudogenes with their corresponding parental genes. Expansion using high-confidence interactions (score ≥ 0.7) from STRING version 12.0 yielded a merged gene list (MGL) consisting of 1,085 genes. This list was used to build a weighted undirected graph – referred to as the Gene Graph (GG) – in which edge weights correspond to STRING PPI scores (Figure 1). Another graph was generated for visualization of shared genes between different EGLs, and it is presented in Figure 2.

**Figure 2.**
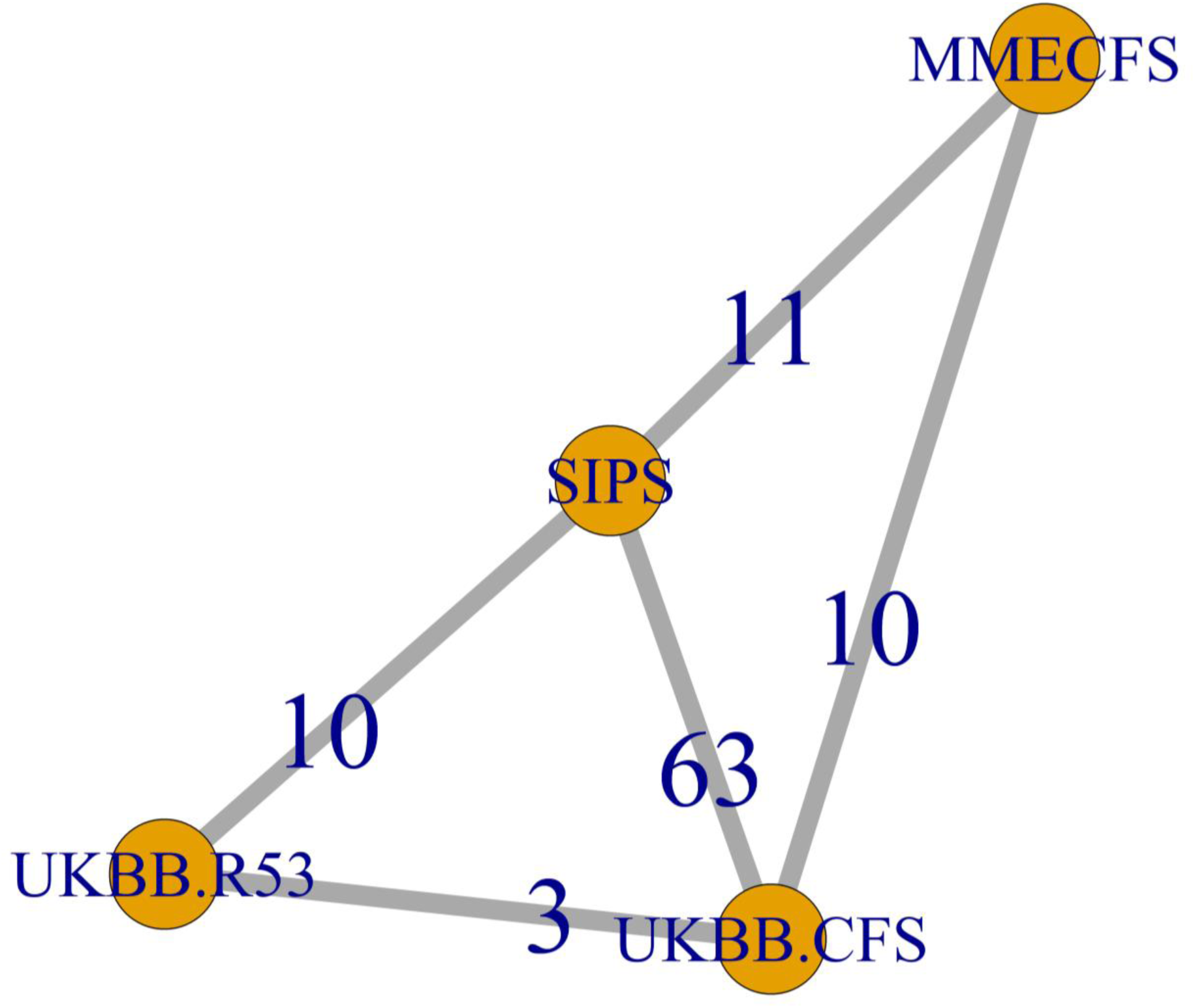
List graph (LG). Weighted undirected graph associated with the four expanded gene lists (EGLs). Each node of the graph corresponds to one of the lists of Table 1 and Table 2. If two EGLs share a number *n* of genes, an edge with weight *n* between the corresponding nodes is drawn. LG is a connected graph. SIPS and UKBB.CFS share genes with each one of the other EGLs, while UKBB.R53 and MMECFS overlap with only two other EGLs each.

Using RWR, we ranked all genes in the MGL based on their connectivity to the seeds. The top 250 candidate genes, together with the original 22 seeds, define the ME/CFS module, whose structure is visualized in Figure 3 and whose gene list is provided in Supplementary Table S7.

**Figure 3.**
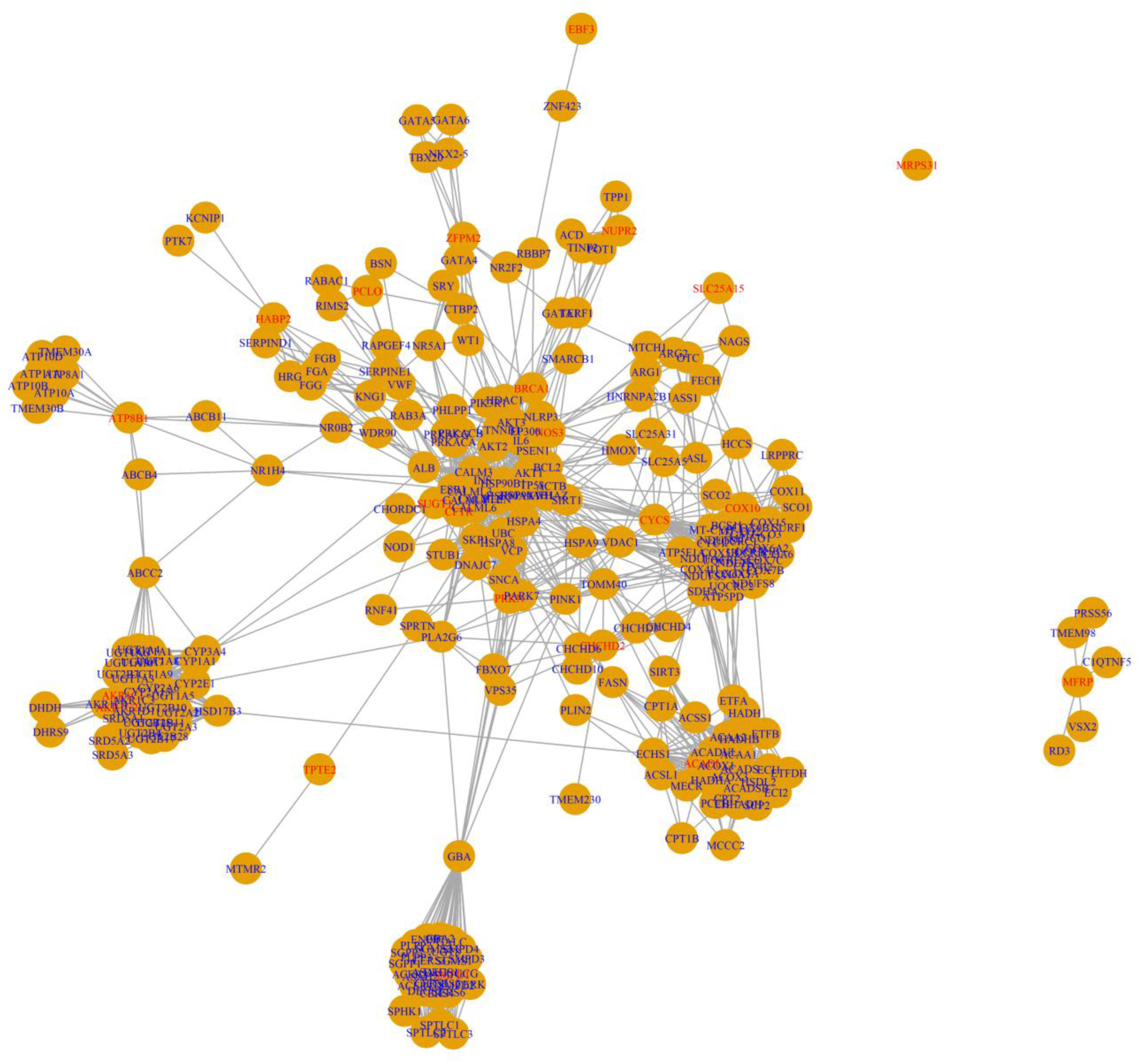
Graph of the ME/CFS module, including the top 250 scoring candidate genes plus the original twenty-two seed genes (in red). An edge between two genes indicates a probability of interaction between them above 70%, according to the STRING database. The number of edges is 2292. The nodes of this graph are selected among the nodes of the graph of Figure 1 by a diffusion method called random walk with restart (RWR), which ranks candidate genes according to their level of connectivity with all the seeds within each connected component of the graph. By comparison with the graph of the complete merged gene list (MGL) of 1085 nodes (Figure 1), we notice that after removing genes with low ranking from RWR, we remain with three connected components, with the largest composed of 265 nodes, one of six nodes (the component composed by seed MFRP of list SIPS), and the last one with only one node (seed MRPS31 of UKBB.CFS). We notice that the main component connects 20 seeds with a contribution from each one of the primary gene lists (PGLs) of Table 1 and Table 2. We also note that the three parental genes (*TPTE2, CYCS*, and *SUGT1*) included as a proxy of their pseudogenes occupy a place in the main component of the ME/CFS module. The genes of the ME/CFS module, with annotations on dosage sensitivity and probability of dominant inheritance, can be found in Supplementary Table S7. The adjacency matrix associated with this graph is in Supplementary Table S9.

To assess whether the ME/CFS gene list represents a biologically meaningful structure rather than a random assembly of genes, we generated 600 MGLs using randomly selected seed genes that mirrored the size and structure of the original PGLs. We then compared the topological features of the experimental ME/CFS MGL with those of the random MGLs using multiple statistical approaches. PCA was performed on six network metrics derived from both GGs and LGs, including connectivity and edge weight. The projection of the ME/CFS MGL fell outside the 95% confidence ellipse in all principal planes (PC1–PC2, PC1–PC3, PC2–PC3), suggesting that its network topology is significantly different from that of randomly constructed gene sets (Figure 4 and Figure S 1). To confirm this result with an orthogonal method, we applied the Local Outlier Factor (LOF) algorithm for anomaly detection. The ME/CFS MGL ranked within the most extreme 5% of LOF values across all random sets (LOF = 1.91), reinforcing the interpretation that the module is not consistent with randomness (Figure 5). The distributions of the variables in the random MGLs are compared with the values of the ME/CFS MGL in Figure 6.

**Figure 4.**
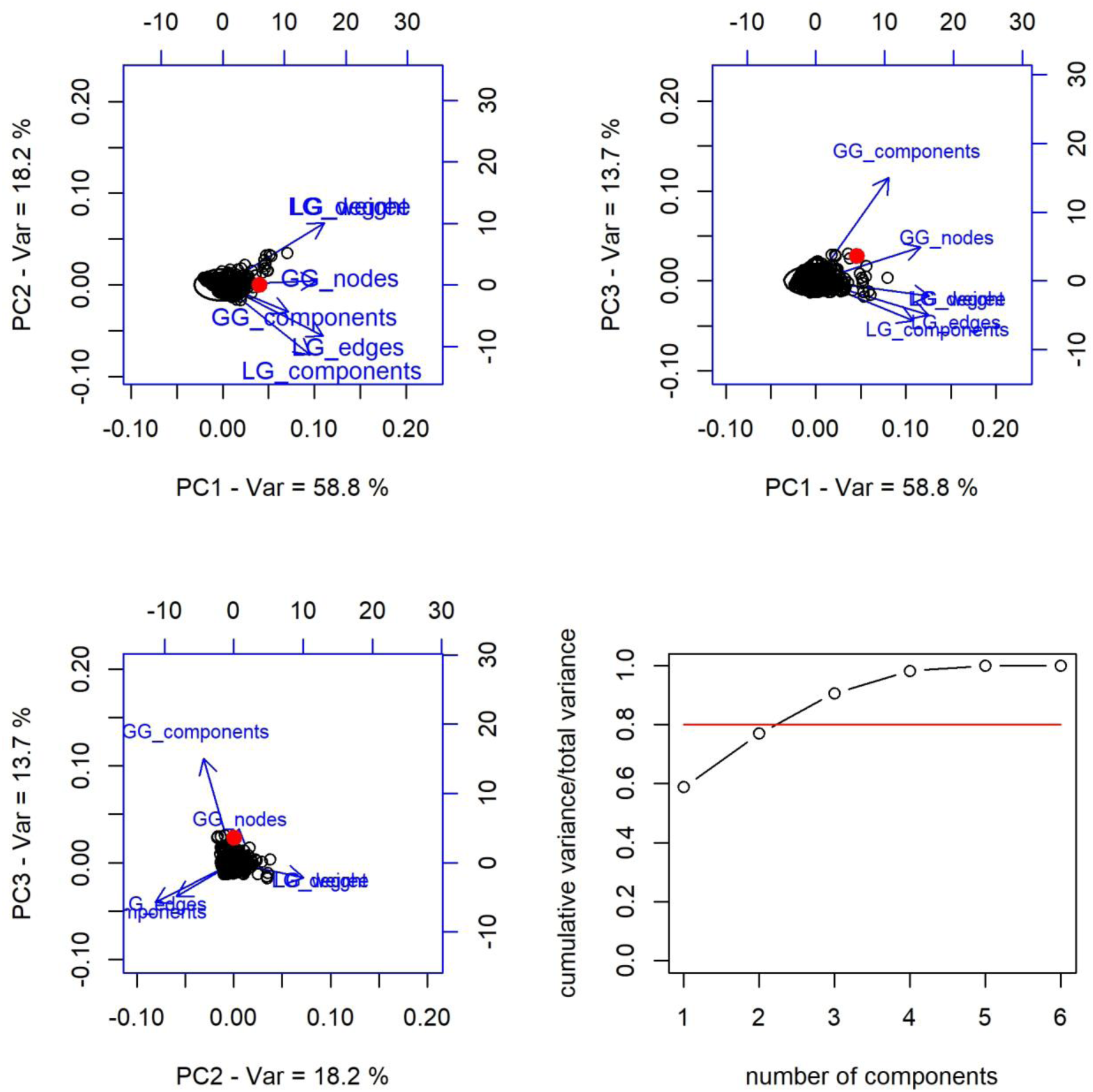
Principal component analysis (PCA) for the comparison between the merged gene list (MGL) built from seeds in Table 1 and Table 2 and 600 MGLs built from random seeds from the human genome. Plane PC1-PC2 (top-left), plane PC1-PC3 (top-right), plane PC2-PC3 (bottom-left), and cumulative variance as a function of the number of principal components (bottom-right). The first two principal components explain about 80% of the total variance. The variables considered are as follows. GG_components: inverse of the number of connected components of the gene graph; GG_nodes: number of nodes of the gene graph; LG_components: inverse of the number of connected components of the list graph; LG_edges: number of edges of the list graph; LG_weight: sum of weights of the edges of the list graph; LG_degree: mean degree of the nodes of the list graph. The MGL from experimental seeds (red dot) falls outside the 95% confidence ellipse on all three principal planes considered. A variant representation of the output of PCA is reported in Figure S 1. The histograms for the values of the variables can be found in Figure 6. This figure was plotted using `biplot()`, an R function.

**Figure 5.**
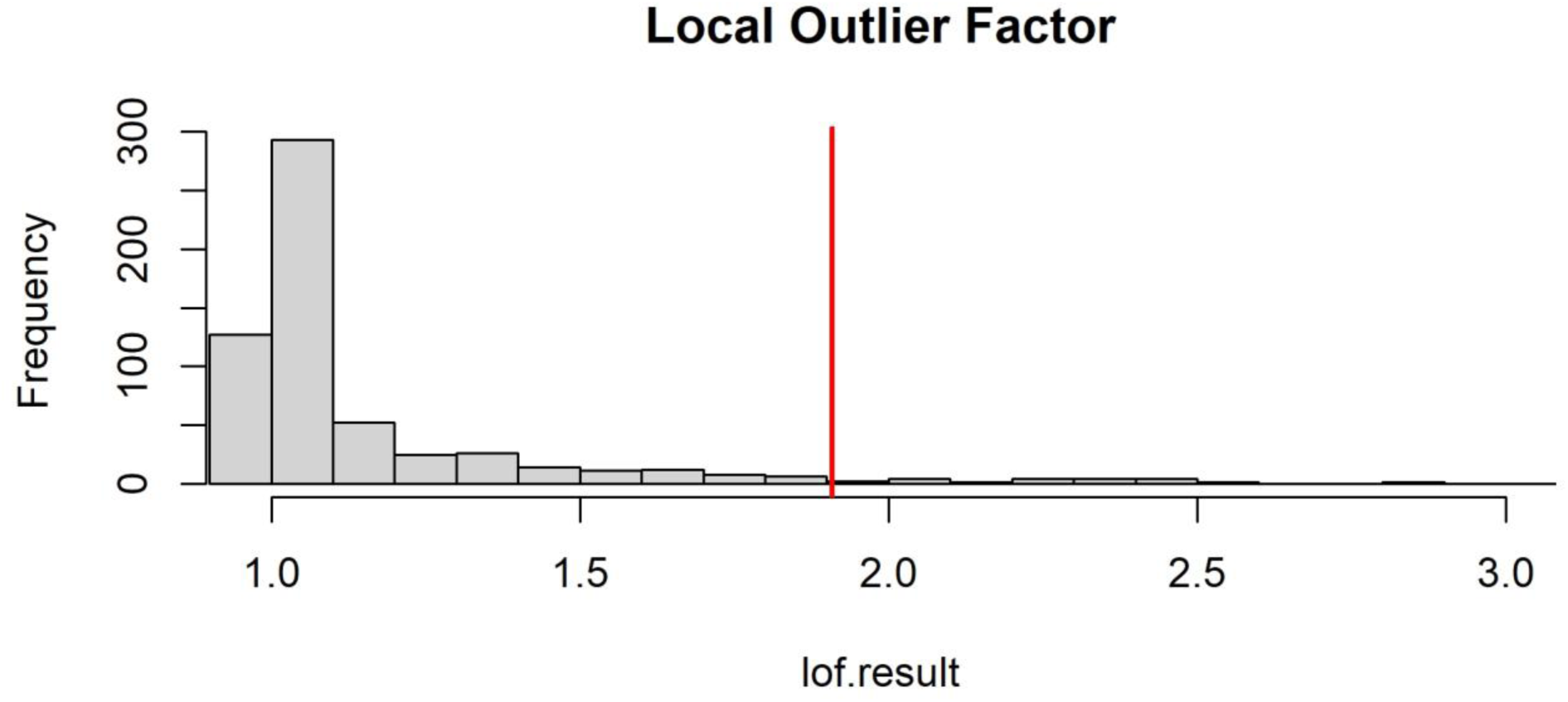
Local outlier factor (LOF) analysis. According to (58) LOF was computed for each one of the random genetic lists, plus the experimental one, as follows: for minPts from 30 to 50, we calculated the maximum LOF of each one of 600 randomly generated merged gene lists (MGLs), and for the one built for ME/CFS. This is the distribution of the LOFs of the total 601 MGLs, and the red line indicates the value corresponding to the ME/CFS MGL. With LOF = 1.91, the MGL of ME/CFS has one of the most extreme 5% LOF. The calculation of LOF was performed with function `lof()` of package *dbscan*.

**Figure 6.**
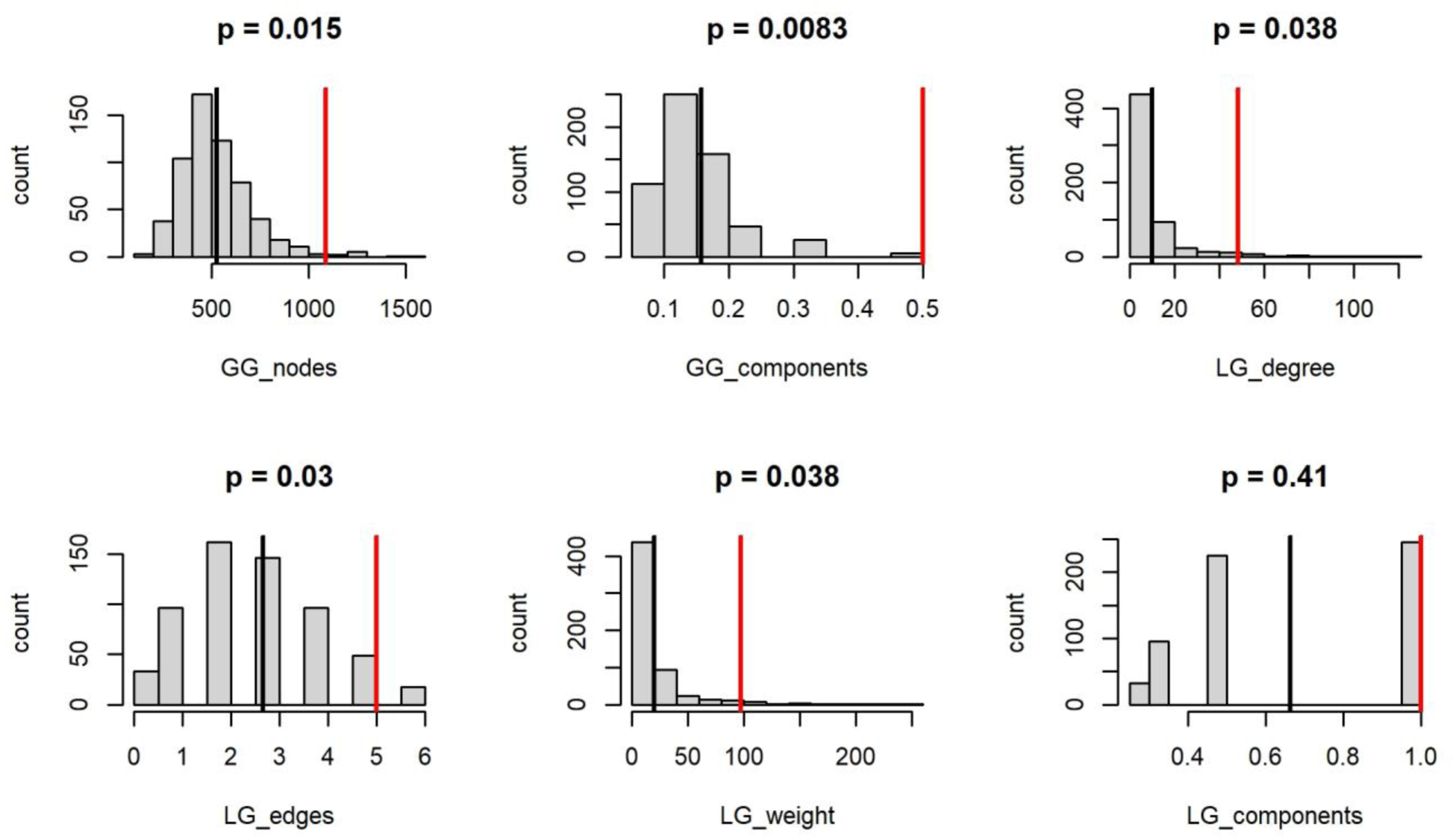
Comparison of the ME/CFS merged gene lists (MGL) (red lines) with six hundred random MGLs built with the same algorithm but using random human seed genes. The variables considered are as follows. GG_components: inverse of the number of connected components of the gene graph; GG_nodes: number of nodes of the gene graph; LG_components: inverse of the number of connected components of the list graph; LG_edges: number of edges of the list graph; LG_weight: total weight of the edges of the list graph; LG_degree: mean degree of the nodes of the list graph. Black lines indicate mean values for random MGLs. Non-parametric, single-tailed statistical testing was employed for *p*-value calculation.

Over-representation analysis using the ME/CFS module on KEGG, Reactome, and DO databases yielded the results shown in Table 9, Table 10, and Table 11. Only terms with contributions from more than two EGLs were retained, and the top 10 results per database were reported. The top 20 terms, along with the overlapping genes, are available in the following supplementary tables: Table S4, Table S5, and Table S6. For visual interpretation, the genes from the ME/CFS module mapped to the top five terms for each database are depicted using chord diagrams in Figure 7, Figure 8, and Figure 9. Four of the top KEGG pathways, with the overlapping genes from the ME/CFS module highlighted in red, are reported in Figure S 2, Figure S 3, Figure S 4, and Figure S 5.

**Figure 7.**
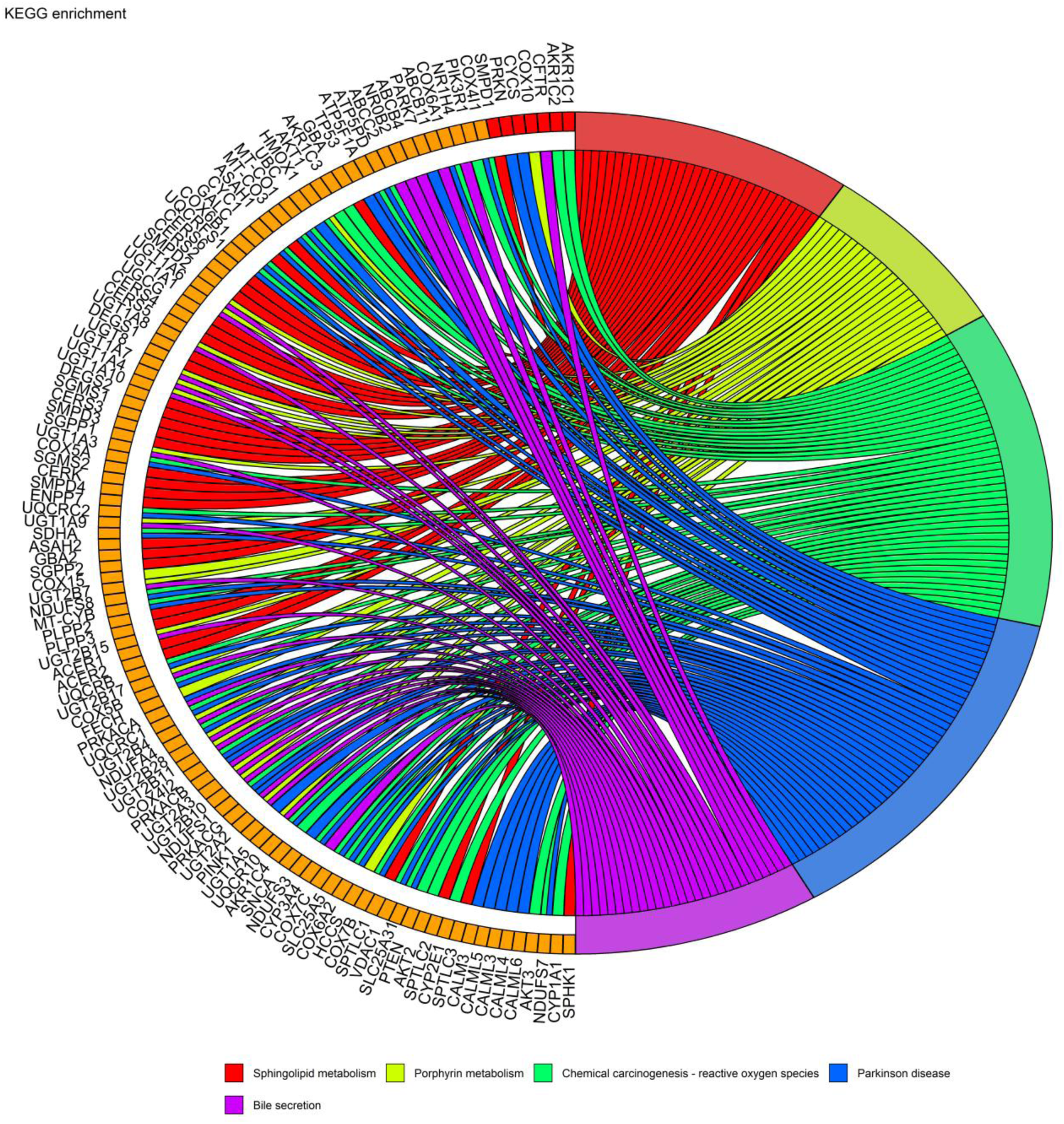
Over-representation analysis of the ME/CFS module (272 genes) on KEGG. Only the top five enriched terms with a contribution from at least two expanded gene lists are considered here (for the other terms, see Table 9 and Supplementary Table S4). In this plot, each term is connected by a ribbon to all the genes of the module that are mapped to it. For each term, a distinct colour is used. For instance, all the genes reached by a red ribbon contribute to the KEGG term *Sphingolipid metabolism*. This figure is generated by function `GOchord()`of Package *GOPlot* (102), with adaptation.

**Figure 8.**
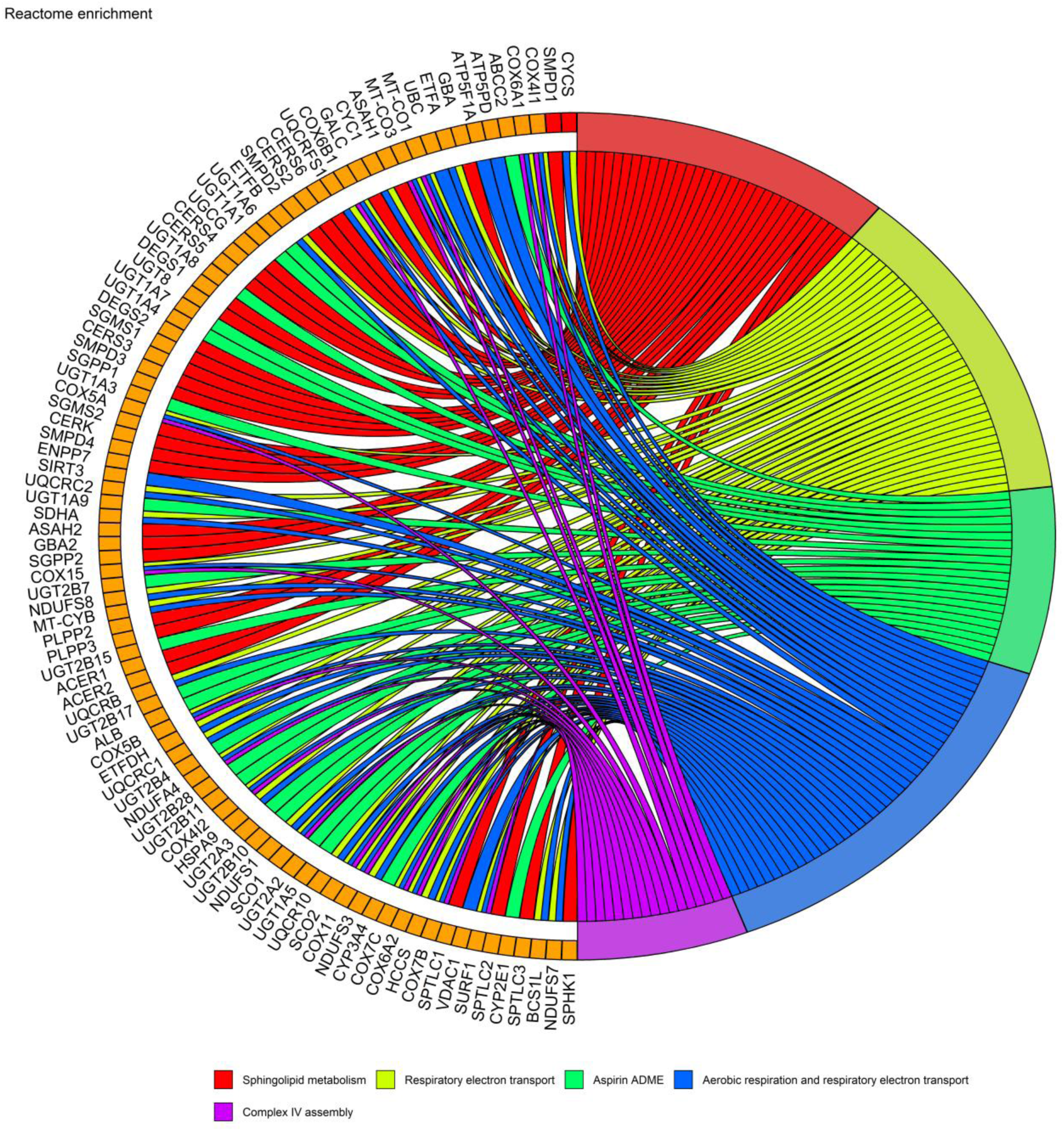
Over-representation analysis of the ME/CFS module (272 genes) on Reactome. Only the top five enriched terms with a contribution from at least two expanded gene lists are considered here (for the other terms, see Table 10 and Supplementary Table S5). In this plot, each term is connected by a ribbon to all the genes of the module that are mapped to it. For each term, a distinct colour is used. For instance, all the genes reached by a red ribbon contribute to the Reactome term *Sphingolipid metabolism*. This figure is generated by function `GOchord()`of Package *GOPlot* (102), with adaptation.

**Figure 9.**
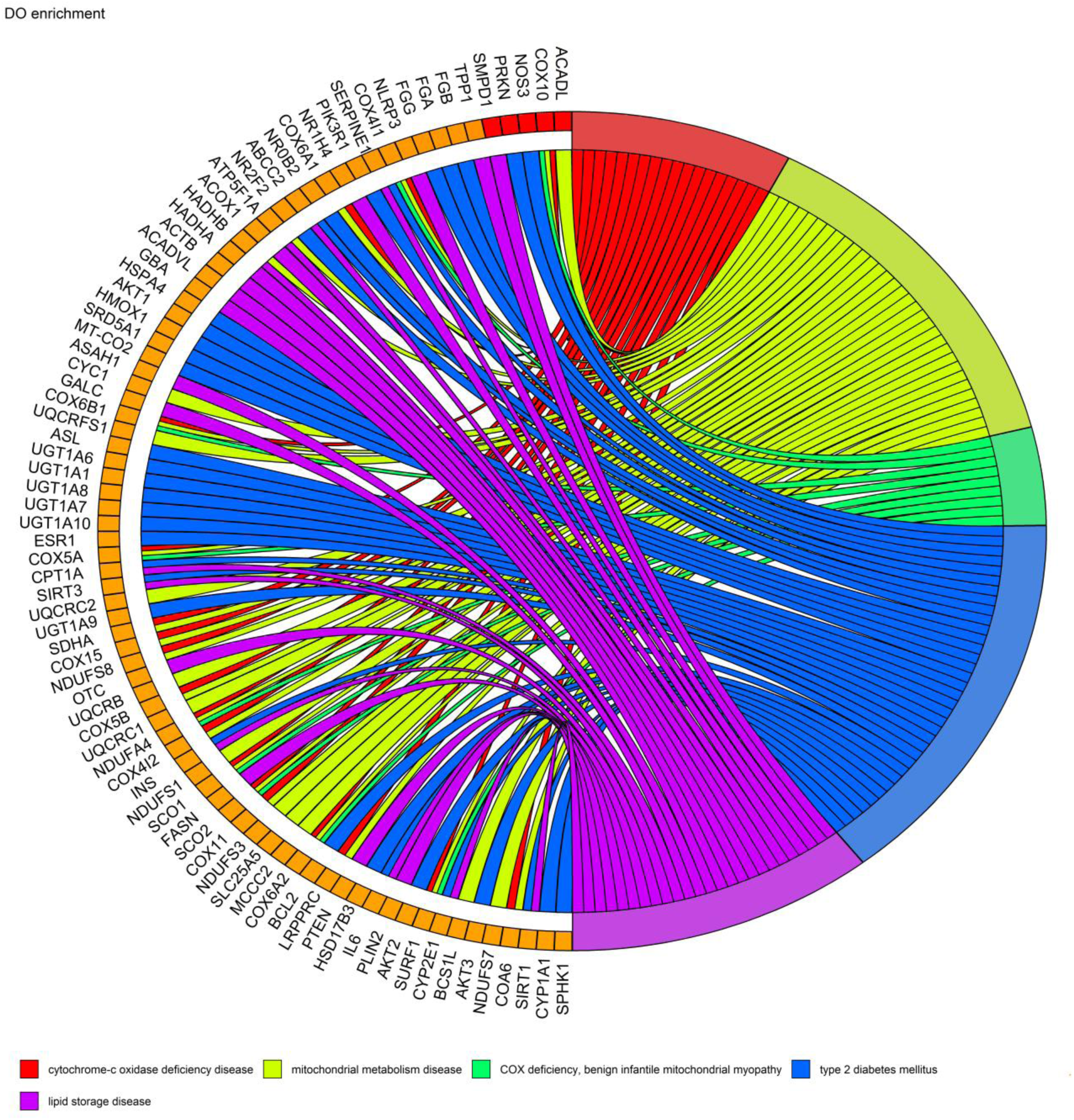
Over-representation analysis of the ME/CFS module (272 genes) on Disease Ontology (DO). Only the top five enriched terms with a contribution from at least two expanded gene lists are considered here (for the other terms, see Table 11 and Supplementary Table S6). In this plot, each term is connected by a ribbon to all the genes of the module that are mapped to it. For each term, a distinct colour is used. For instance, all the genes reached by a red ribbon contribute to the DO term *Cytochrome C deficiency disease*. This figure is generated by function `GOchord()`of Package *GOPlot* (102), with adaptation.

**Table 9.**
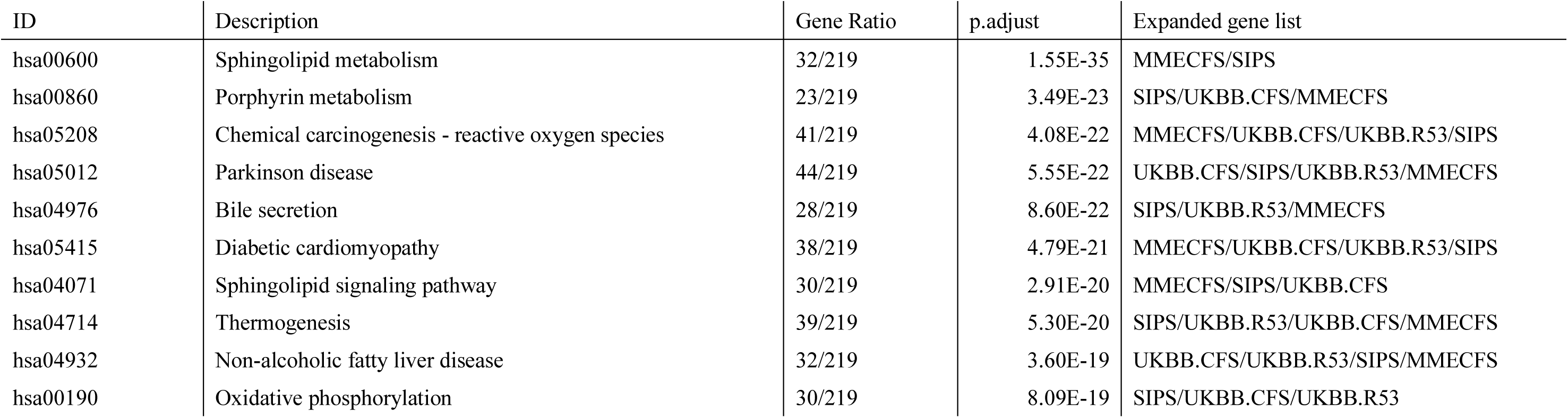
Over-representation analysis (ORA) on KEGG of the 272 genes of the ME/CFS module. ORA was performed by function `enrichKEGG()`of package *ClusterProfile* (see Table 7). Results with a contribution of only one primary gene list were excluded, and only the top ten terms were included. A list of the top twenty results, with the genes overlapping each term, is included in Supplementary Table S4. The genes overlapping with the first five terms of the list are shown in Figure 7. Pathways for sphingolipid metabolism (hsa00600), porphyrin metabolism (hsa00860), Parkinson’s disease (hsa05012), and sphingolipid signaling (hsa04071) – with the overlapping genes from the ME/CFS module highlighted – can be found in Figure S 3, Figure S 4, Figure S 5, and Figure S 6, respectively.

| ID | Description | Gene Ratio | p.adjust | Expanded gene list |
| --- | --- | --- | --- | --- |
| hsa00600 | Sphingolipid metabolism | 32/219 | 1.55E-35 | MMECFS/SIPS |
| hsa00860 | Porphyrin metabolism | 23/219 | 3.49E-23 | SIPS/UKBB.CFS/MMECFS |
| hsa05208 | Chemical carcinogenesis - reactive oxygen species | 41/219 | 4.08E-22 | MMECFS/UKBB.CFS/UKBB.R53/SIPS |
| hsa05012 | Parkinson disease | 44/219 | 5.55E-22 | UKBB.CFS/SIPS/UKBB.R53/MMECFS |
| hsa04976 | Bile secretion | 28/219 | 8.60E-22 | SIPS/UKBB.R53/MMECFS |
| hsa05415 | Diabetic cardiomyopathy | 38/219 | 4.79E-21 | MMECFS/UKBB.CFS/UKBB.R53/SIPS |
| hsa04071 | Sphingolipid signaling pathway | 30/219 | 2.91E-20 | MMECFS/SIPS/UKBB.CFS |
| hsa04714 | Thermogenesis | 39/219 | 5.30E-20 | SIPS/UKBB.R53/UKBB.CFS/MMECFS |
| hsa04932 | Non-alcoholic fatty liver disease | 32/219 | 3.60E-19 | UKBB.CFS/UKBB.R53/SIPS/MMECFS |
| hsa00190 | Oxidative phosphorylation | 30/219 | 8.09E-19 | SIPS/UKBB.CFS/UKBB.R53 |

**Table 10.** Over-representation analysis (ORA) on Reactome of the 272 genes of the ME/CFS module. ORA was performed by function `enrichPA()`of package *ReactomePA* (see Table 7). Results with a contribution of only one primary gene list were excluded, and only the top ten terms were included. A list of the top twenty results, with the genes overlapping each term, is included in Supplementary Table S5. The genes overlapping with the first five terms of the list are shown in Figure 8.

| ID | Description | Gene Ratio | p.adjust | Expanded gene list |
| --- | --- | --- | --- | --- |
| R-HSA-428157 | Sphingolipid metabolism | 32/248 | 5.50E-25 | MMECFs/SIPS |
| R-HSA-611105 | Respiratory electron transport | 36/248 | 6.03E-24 | UKBB.CFS/UKBB.R53/SIPS |
| R-HSA-9749641 | Aspirin ADME | 21/248 | 2.19E-21 | UKBB.R53/MMECFs/UKBB.CFS |
| R-HSA-1428517 | Aerobic respiration and respiratory electron transport | 41/248 | 2.19E-21 | UKBB.CFS/UKBB.R53/SIPS |
| R-HSA-9864848 | Complex IV assembly | 17/248 | 1.81E-14 | UKBB.CFS/UKBB.R53/SIPS |
| R-HSA-9748784 | Drug ADME | 23/248 | 2.67E-14 | MMECFs/UKBB.R53/UKBB.CFS |
| R-HSA-5628897 | TP53 Regulates Metabolic Genes | 20/248 | 1.58E-13 | UKBB.CFS/UKBB.R53/SIPS/MMECFs |
| R-HSA-8978868 | Fatty acid metabolism | 25/248 | 1.24E-11 | SIPS/MMECFs |
| R-HSA-9707564 | Cytoprotection by HMOX1 | 16/248 | 1.42E-11 | UKBB.CFS/UKBB.R53/SIPS/MMECFs |
| R-HSA-9837999 | Mitochondrial protein degradation | 19/248 | 3.34E-11 | UKBB.R53/UKBB.CFS/SIPS/MMECFs |

**Table 11.** Over-representation analysis (ORA) on Disease Ontology (DO) of the 272 genes of the ME/CFS module. ORA was performed by function `enrichDO()`of package *DOSE* (see Table 7). Results with a contribution of only one primary gene list were excluded, and only the top ten terms were included. A list of the top twenty results, with the genes overlapping each term, is included in Supplementary Table S6. The genes overlapping with the first five terms of the list are shown in Figure 9.

| ID | Description | Gene Ratio | p.adjust | Expanded gene list |
| --- | --- | --- | --- | --- |
| DOID:3762 | cytochrome-c oxidase deficiency disease | 18/212 | 6.72E-15 | SIPS/UKBB.CFS/UKBB.R53 |
| DOID:700 | mitochondrial metabolism disease | 32/212 | 7.79E-13 | SIPS/UKBB.CFS/UKBB.R53 |
| DOID:0081377 | COX deficiency, benign infantile mitochondrial myopathy | 9/212 | 6.08E-08 | SIPS/UKBB.CFS/UKBB.R53 |
| DOID:9352 | type 2 diabetes mellitus | 35/212 | 1.74E-06 | MMECFs/SIPS/UKBB.R53/UKBB.CFS |
| DOID:9455 | lipid storage disease | 25/212 | 4.13E-06 | MMECFs/UKBB.R53/UKBB.CFS/SIPS |
| DOID:9267 | urea cycle disorder | 6/212 | 1.54E-05 | UKBB.CFS/MMECFs |
| DOID:9452 | steatotic liver disease | 20/212 | 2.58E-05 | UKBB.CFS/SIPS/UKBB.R53/MMECFs |
| DOID:3211 | lysosomal storage disease | 25/212 | 2.58E-05 | MMECFs/UKBB.R53/UKBB.CFS/SIPS |
| DOID:1247 | blood coagulation disease | 15/212 | 2.73E-05 | UKBB.CFS/SIPS/MMECFs |
| DOID:10652 | Alzheimer's disease | 30/212 | 2.73E-05 | SIPS/MMECFs/UKBB.CFS/UKBB.R53 |

The ME/CFS module overlaps with sphingolipid metabolism and signaling, heme degradation, and oxidative phosphorylation on both the KEGG and Reactome databases. KEGG also highlighted the over-representation of genes involved in xenobiotic metabolism and the cellular response to reactive oxygen species, and genes involved in thermogenesis. Reactome results pointed to drug metabolism and excretion, as well as two additional energy-related pathways: fatty acid metabolism and metabolic regulation by *TP53*. Both KEGG and Disease Ontology revealed an overlap with neurodegenerative disorders and steatotic liver disease (Table 9, Table 10, and Table 11).

Tissue-specific enrichment analysis using the Human Protein Atlas revealed over-representation of genes specifically expressed in the liver, the small intestine, and gallbladder (Figure S 7).

These findings suggest an involvement of lipid metabolism, mitochondrial respiratory function, and oxidative stress in ME/CFS pathophysiology. The enrichment in genes specifically expressed in the digestive system suggests that gastrointestinal and hepatobiliary tissues may be central targets in disease development.

The probabilities of haploinsufficiency and triplosensitivity for the 250 top-ranking candidate genes plus the 22 seed genes are significantly greater than what we find in the complete set of human protein-coding genes (Table 5). This enrichment suggests that structural variants, particularly copy number variants, may play a pathogenic role in some cases of Mendelian ME/CFS.

## 4 Discussion

In this study, we selected twenty-two genes with experimental evidence supporting a possible involvement in the pathogenesis of ME/CFS and used them as seeds to identify candidate genes with high-confidence interactions, based on the STRING protein–protein interaction database. To prioritize these candidates, we applied Random Walk with Restart (RWR), a graph-based algorithm that ranks genes by their network proximity to a set of input nodes (seeds). It is well established that genes associated with the same disease tend to be clustered within protein interaction networks, even if no direct interactions exist between them (Goh et al., 2007; Oti et al., 2006; Xu & Li, 2006). RWR has been shown to successfully prioritize disease-relevant genes from large candidate pools (Cowen et al., 2017; Köhler et al., 2008). Based on this approach, we selected the top 250 ranked genes together with the original 22 seeds to define the ME/CFS module. This curated gene set was then subjected to over-representation analysis (ORA) using the KEGG, Reactome, and Disease Ontology databases to uncover shared biological functions and disease associations.

One of the most significant results of the enrichment analysis performed on the ME/CFS module is sphingolipid metabolism and signaling, identified by ORA on KEGG (Table 9, pathways hsa0600 and hsa04071, Figure S 3, Figure S 6) and Reactome (Table 10). A significant reduction in ceramides, sphingomyelins, and glycosphingolipids was documented in 45 subjects with ME/CFS by targeted metabolomics in plasma (4). Consistent with this, a reduction in sphingomyelins and ceramides in plasma was reported in a cohort of 106 subjects with ME/CFS (63). Recently, an analysis of 249 metabolic biomarkers (168 absolute measures and 81 ratios) measured in plasma by high-throughput NMR in the UKBiobank was performed on a selection of 1194 patients with self-reported ME/CFS, and a reduction in sphingomyelins was among the statistically significant differences with controls (64). Other studies reported fold changes in the opposite direction. A group – using a metabolomic panel of about 1750 compounds in 52 female patients – found that ME/CFS subjects had elevated ceramide and sphingomyelin levels in plasma (65). A Norwegian study profiled serum from 83 patients using both global metabolomics and targeted lipidomics and reported a significant increase in two sphingomyelins and one ceramide in serum (66). A subsequent study confirmed a rise in plasma ceramides and sphingomyelins in 149 ME/CFS patients by untargeted metabolomics (67). Targeted lipidomic analysis reported a rise in five sphingomyelins in cerebrospinal fluid (CSF) in 59 ME/CFS patients (68).

Sphingolipids are lipids that localize predominantly to the plasma membrane, where they regulate key cellular processes, including adhesion, proliferation, migration, and apoptosis. They have been involved in inflammation, neurodegeneration, cancer, and lysosomal storage disorders (69). Ceramides have been implicated in the regulation of cellular bioenergetics (70). In the context of ME/CFS, the meaning of alterations in sphingolipid metabolism in blood has been variously interpreted as the manifestation of a broad hypometabolic shift (4), or a downstream effect of the pathological process and the lack of physical activity (65). Baraniuk interpreted his findings of increased sphingomyelins in CSF as a possible sign of myelin and white matter dysfunction (68).

Enrichment analysis on KEGG selected porphyrin metabolism as the second most significant term (Table 9). The overlap between this pathway and the ME/CFS module maps specifically to the branch involved in heme catabolism that leads to the formation of bilirubin, biliverdin and related metabolites (Figure S 4). This finding is supported by Reactome, where the term *Cytoprotection by HMOXI* – referring to the degradation of heme into biliverdin – is also significantly enriched (Table 10). Perturbations in this segment of the human metabolism have previously been documented in ME/CFS. Two independent studies using UK Biobank data reported reductions in both total and direct bilirubin in ME/CFS patients: one in a cohort of 1,455 individuals (14) and the other in a partially overlapping group of 1,194 patients (64). In a previous study on 32 females with ME/CFS, one of the top findings was elevated blood heme in patients (6). The heme group – a heterocyclic ring with an iron at its centre – is a porphyrin derivative contained by myoglobin, haemoglobin, cytochromes such as cytochrome P450 and Complex IV, and other enzymes. The catabolism of heme is considered part of the antioxidant defence (71).

The third most significantly enriched KEGG term is associated with mitochondrial production of reactive oxygen species (ROS) and the metabolism of xenobiotics, including their non-genotoxic carcinogenic effects (Table 9). Several markers of redox imbalance that correlate with the severity of symptoms have been reported in ME/CFS (72). Furthermore, ME/CFS is a significant risk factor for developing non-Hodgkin lymphoma (NHL) (73), with a recent metanalysis showing that chemical exposure is a significant risk factor for NHL (74).

Reactome found an overlap with drug Absorption, Distribution, Metabolism, and Excretion (ADME). To our knowledge, there is a lack of formal investigation into drug ADME among subjects with ME/CFS, but some anecdotal evidence suggests that people with ME/CFS are often particularly sensitive to medications (75).

An overlap between the ME/CFS module and neurodegenerative diseases, namely Parkinson’s disease and Alzheimer’s disease, is suggested by KEGG and DO, respectively (Table 9, Table 11, Figure S 5). A similar result was obtained in a study using a set of seeds partially overlapping with the one used by us but with a different pipeline (25). Abnormalities in the basal ganglia and the midbrain – anatomical regions associated with PD’s etiology – have been documented in ME/CFS (8, 15, 76, 77) and mitochondrial dysfunction has been implicated in the loss of dopaminergic neurons in PD (78) and reported in ME/CFS (79, 80).

Bile acid metabolism – identified by KEGG as significantly overlapping with the ME/CFS module – was implicated in ME/CFS by some metabolomic studies, reporting a depletion in bile acids (4, 5, 67). Bile acids are essential for the digestion and absorption of dietary lipids, therefore, this result refers, more broadly, to lipid metabolism.

In our analysis, the Reactome term *TP53 regulates metabolic genes* emerged as significantly enriched (Table 10). This pathway centres on the transcriptional regulatory functions of *TP53*, a tumour suppressor traditionally known for its roles in cell cycle control and apoptosis (81, 82) but increasingly recognized as a key metabolic regulator. The protein encoded by *TP53* modulates cellular energy metabolism by suppressing glycolysis and promoting oxidative phosphorylation and fatty acid oxidation, as reviewed in (83), where the hypothesis of its involvement in ME/CFS was formulated. For instance, in a study on 18 carriers of gain of function mutations on *TP53*, increased mitochondrial function is reported (84). A longitudinal cytokine profiling conducted on an exceptionally severe ME/CFS patient, a male with hypermobility spectrum disorder (HSD), revealed *TP53* activation during a period of substantial improvement (85). In another study, a rare heterozygous *TP53* mutation (c.455C>T, p.P152.L) was reported in a woman with a ME/CFS diagnosis who developed worsening fatigue and exercise intolerance after an episode of mononucleosis at age 16. Interestingly, this patient showed reduced mitochondrial efficiency, but a series of experiments linked to WASF3 overexpression, rather than to direct consequence of the mutation (80).

One term on KEGG, four on Reactome, and three on Disease Ontology refer to mitochondrial abnormalities, particularly involving the electron transport chain. On Reactome, ORA also detected a significant overlap with fatty acid metabolism, another pathway in cellular energy production. In ME/CFS, we have studies reporting mitochondrial structural and functional abnormalities, albeit with some inconsistencies (79), studies documenting impaired aerobic performances using a 2-day CPET methodology (11, 12), and blood biomarkers of reduced energy metabolism, particularly involving the tricarboxylic acid (TCA) and fatty acid oxidation (4–7).

The ME/CFS module is enriched for genes expressed in the liver, small intestine, and gallbladder (Figure S 7). Elevated levels of hepatic enzymes (AST, ALT, and GGT) have been documented among the ME/CFS patients within the UKBiobank cohort (14). This finding, along with the already mentioned reduction in bilirubin and bile acids, may suggest some form of liver dysfunction in this population. Gastrointestinal symptoms, like irritable bowel syndrome (IBS), are included in the International Consensus Criteria (86), one of the diagnostic tools for ME/CFS.

This analysis did not find a direct link to immunity; therefore, according to our results, ME/CFS immune dysregulation (10, 13, 15) may be a downstream process.

Our study highlights an overlap of the ME/CFS module with lipid metabolism – sphingolipid and fatty acid metabolism in particular – and with energy metabolism, particularly oxidative phosphorylation. Enrichment was also observed for genes involved in heme degradation and metabolic regulation by *TP53*. An overlap with neurodegenerative and metabolic diseases is suggested. The heterogeneity of these results may reflect either noise in the seeds we selected or the heterogeneous nature of the nosological entity we are trying to define. Nevertheless, we note that the findings of the present analysis partially overlap with a disease model for ME/CFS proposed in (72), in which redox imbalance, mitochondrial dysfunction, and inflammation are engaged in pairwise bidirectional feedback loops. In this model, an initial disruption in any of these three domains can initiate a self-sustaining pathological state. By adopting this model, a unifying interpretation of our results may be that impaired mitochondrial respiration and disrupted heme catabolism contribute to oxidative stress, which in turn exacerbates mitochondrial impairment in a vicious cycle. Mitochondrial impairment could stem from intrinsic defects in respiratory complexes or arise secondarily through aberrant ceramide signaling, deficient fatty acid oxidation, or dysregulation of the *TP53* metabolic control axis. These mechanisms may be particularly relevant in the liver and small intestine, as suggested by tissue-specific gene expression enrichment. Enrichment in the thermogenesis pathway (Table 9) raises the possibility that excessive reactive oxygen species (ROS) generation during fat oxidation in brown or beige adipocytes could contribute to disease pathology. This aligns with the reported symptoms of temperature instability and cold intolerance in ME/CFS (86).

The ME/CFS module (Table S7) may serve as a tool for studying Mendelian forms of ME/CFS. While thousands of rare genetic diseases have already been described, theoretical estimates suggest many remain undiscovered (87). New rare genetic diseases have been recently identified among patients previously collected in umbrella clinical designations like autism, intellectual disability, and congenital heart failure (87). As in the case of cognitive deficits, unexplained fatigue and exercise intolerance may be caused by different mechanisms, leaving room not only for more than one single type of idiopathic ME/CFS but also for different monogenic forms. While curated gene panels have aided diagnosis in many rare diseases (88, 89), no such tool yet exists for ME/CFS. This study offers a first iteration, integrating published evidence and known protein–protein interactions, but new experimental data from both GWAS-type studies and NGS analyses are urgently needed.

We note that in a pipeline like the one described here, where a list expansion procedure is applied to seeds and then enrichment analysis is performed, results can be due to chance alone since Reactome and KEGG are among the resources employed for score calculation in STRING. The ME/CFS module has been selected from an MGL with characteristics uncommon in randomly generated MGLs, nevertheless, we acknowledge the risk that our results are partly due to the algorithm that generated the module rather than biological information contained in the seeds. Another weakness of this study is the substitution of three pseudogenes with their respective parental genes in the network analysis. We also acknowledge as a limitation the inclusion of a cohort of patients with chronic fatigue (R53), a condition only related to ME/CFS. Moreover, the ME/CFS cohort from the UK Biobank database was self-reported.

## 5 Conclusion

This study presents a systematic attempt to define a disease module for ME/CFS based on current genetic data and protein interaction networks. By integrating findings from a population-level study and case reports, we generated a ranked gene list that showed an overlap with biological processes implicated in ME/CFS, including sphingolipid metabolism, mitochondrial function, and heme catabolism. The ME/CFS module reflects known metabolic disturbances observed in patient cohorts and highlights novel candidate pathways, such as TP53-regulated metabolic genes and thermogenesis. This gene list is proposed as a first draft toward a diagnostic aid for Mendelian ME/CFS and is expected to evolve with new data from whole-genome sequencing and future GWAS studies.

## Data Availability

All data produced in the present work are contained in the manuscript and supplementary materials

## References

1. Chu L, Valencia IJ, Garvert DW, Montoya JG. Onset Patterns and Course of Myalgic Encephalomyelitis/Chronic Fatigue Syndrome. Front Pediatr. 2019;7:12.

2. IOM. The National Academies Collection: Reports funded by National Institutes of Health. Beyond Myalgic Encephalomyelitis/Chronic Fatigue Syndrome: Redefining an Illness. Washington (DC): National Academies Press (US) Copyright 2015 by the National Academy of Sciences. All rights reserved.; 2015.

3. Komaroff AL. Advances in Understanding the Pathophysiology of Chronic Fatigue Syndrome. JAMA. 2019.

4. Naviaux RK, Naviaux JC, Li K, Bright AT, Alaynick WA, Wang L, et al. Metabolic features of chronic fatigue syndrome. Proc Natl Acad Sci U S A. 2016;113(37):E5472–80.

5. Germain A, Ruppert D, Levine SM, Hanson MR. Metabolic profiling of a myalgic encephalomyelitis/chronic fatigue syndrome discovery cohort reveals disturbances in fatty acid and lipid metabolism. Mol Biosyst. 2017;13(2):371–9.

6. Germain A, Ruppert D, Levine SM, Hanson MR. Prospective Biomarkers from Plasma Metabolomics of Myalgic Encephalomyelitis/Chronic Fatigue Syndrome Implicate Redox Imbalance in Disease Symptomatology. Metabolites. 2018;8(4).

7. Germain A, Giloteaux L, Moore GE, Levine SM, Chia JK, Keller BA, et al. Plasma metabolomics reveals disrupted response and recovery following maximal exercise in myalgic encephalomyelitis/chronic fatigue syndrome. JCI Insight. 2022;7(9).

8. Tirelli U, Chierichetti F, Tavio M, Simonelli C, Bianchin G, Zanco P, et al. Brain positron emission tomography (PET) in chronic fatigue syndrome: preliminary data. Am J Med. 1998;105(3A):54S–8S.

9. Siessmeier T, Nix WA, Hardt J, Schreckenberger M, Egle UT, Bartenstein P. Observer independent analysis of cerebral glucose metabolism in patients with chronic fatigue syndrome. J Neurol Neurosurg Psychiatry. 2003;74(7):922–8.

10. Mueller C, Lin JC, Sheriff S, Maudsley AA, Younger JW. Evidence of widespread metabolite abnormalities in Myalgic encephalomyelitis/chronic fatigue syndrome: assessment with whole-brain magnetic resonance spectroscopy. Brain Imaging Behav. 2019.

11. Stevens S, Snell C, Stevens J, Keller B, VanNess JM. Cardiopulmonary Exercise Test Methodology for Assessing Exertion Intolerance in Myalgic Encephalomyelitis/Chronic Fatigue Syndrome. Frontiers in Pediatrics. 2018;6:242.

12. Franklin JD, Atkinson G, Atkinson JM, Batterham AM. Peak Oxygen Uptake in Chronic Fatigue Syndrome/Myalgic Encephalomyelitis: A Meta-Analysis. Int J Sports Med. 2019;40(2):77–87.

13. Montoya JG, Holmes TH, Anderson JN, Maecker HT, Rosenberg-Hasson Y, Valencia IJ, et al. Cytokine signature associated with disease severity in chronic fatigue syndrome patients. Proc Natl Acad Sci U S A. 2017;114(34):E7150–E8.

14. Beentjes SV, Kaczmarczyk J, Cassar A, Samms GL, Hejazi NS, Khamseh A, et al. Replicated blood-based biomarkers for Myalgic Encephalomyelitis not explicable by inactivity. medRxiv. 2024:2024.08.26.24312606.

15. Nakatomi Y, Mizuno K, Ishii A, Wada Y, Tanaka M, Tazawa S, et al. Neuroinflammation in Patients with Chronic Fatigue Syndrome/Myalgic Encephalomyelitis: An ¹¹C-(R)-PK11195 PET Study. J Nucl Med. 2014;55(6):945–50.

16. Lim EJ, Ahn YC, Jang ES, Lee SW, Lee SH, Son CG. Systematic review and meta-analysis of the prevalence of chronic fatigue syndrome/myalgic encephalomyelitis (CFS/ME). J Transl Med. 2020;18(1):100.

17. Bakken IJ, Tveito K, Gunnes N, Ghaderi S, Stoltenberg C, Trogstad L, et al. Two age peaks in the incidence of chronic fatigue syndrome/myalgic encephalomyelitis: a population-based registry study from Norway 2008-2012. BMC Med. 2014;12:167.

18. Kielland A, Liu J. What can wage development before and after a G93.3 diagnosis tell us about prognoses for myalgic encephalomyelitis? Social Sciences & Humanities Open. 2025;11:101206.

19. Albright F, Light K, Light A, Bateman L, Cannon-Albright LA. Evidence for a heritable predisposition to Chronic Fatigue Syndrome. BMC Neurol. 2011;11:62.

20. Walsh CM, Zainal NZ, Middleton SJ, Paykel ES. A family history study of chronic fatigue syndrome. Psychiatric Genetics. 2001;11(3).

21. Underhill RA, O’Gorman R. Prevalence of Chronic Fatigue Syndrome and Chronic Fatigue Within Families of CFS Patients. Journal Of Chronic Fatigue Syndrome. 2006;13(1):3–13.

22. Sudlow C, Gallacher J, Allen N, Beral V, Burton P, Danesh J, et al. UK biobank: an open access resource for identifying the causes of a wide range of complex diseases of middle and old age. PLoS Med. 2015;12(3):e1001779.

23. Dibble JJ, McGrath SJ, Ponting CP. Genetic risk factors of ME/CFS: a critical review. Hum Mol Genet. 2020;29(R1):R117–R24.

24. Neale L. UK Biobank Round 2 GWAS Results (Released 1st August 2018). 2018.

25. Hung L-Y, Wu C-S, Chang C-J, Li P, Hicks K, Dibble JJ, et al. A network medicine approach to investigating ME/CFS pathogenesis in severely ill patients: a pilot study. Frontiers in Human Neuroscience. 2025;19.

26. McGarrity S, Ziehr DR, Austin-Tse CA, Wein MN, Chivukula RR, Oldham WM. Exercise Intolerance and Low Cardiac Filling Pressures in a Woman With a Novel eNOS Mutation. Circulation: Genomic and Precision Medicine. 2024;17(5):e004741.

27. Oakley J, Hill M, Giess A, Tanguy M, Elgar G. Long read sequencing characterises a novel structural variant, revealing underactive AKR1C1 with overactive AKR1C2 as a possible cause of severe chronic fatigue. Journal of Translational Medicine. 2023;21(1):825.

28. Jahanbani F, Maynard RD, Sing JC, Jahanbani S, Perrino JJ, Spacek DV, et al. Phenotypic characteristics of peripheral immune cells of Myalgic encephalomyelitis/chronic fatigue syndrome via transmission electron microscopy: A pilot study. PLoS One. 2022;17(8):e0272703.

29. Szklarczyk D, Kirsch R, Koutrouli M, Nastou K, Mehryary F, Hachilif R, et al. The STRING database in 2023: protein-protein association networks and functional enrichment analyses for any sequenced genome of interest. Nucleic Acids Res. 2023;51(D1):D638–d46.

30. Xu J, Li Y. Discovering disease-genes by topological features in human protein-protein interaction network. Bioinformatics. 2006;22(22):2800–5.

31. Goh KI, Cusick ME, Valle D, Childs B, Vidal M, Barabási AL. The human disease network. Proc Natl Acad Sci U S A. 2007;104(21):8685–90.

32. Oti M, Snel B, Huynen MA, Brunner HG. Predicting disease genes using protein-protein interactions. J Med Genet. 43. England2006. p. 691–8.

33. Barabási AL, Gulbahce N, Loscalzo J. Network medicine: a network-based approach to human disease. Nat Rev Genet. 2011;12(1):56–68.

34. Peltonen L, Perola M, Naukkarinen J, Palotie A. Lessons from studying monogenic disease for common disease. Hum Mol Genet. 2006;15 Spec No 1:R67-74.

35. Freund MK, Burch KS, Shi H, Mancuso N, Kichaev G, Garske KM, et al. Phenotype-Specific Enrichment of Mendelian Disorder Genes near GWAS Regions across 62 Complex Traits. Am J Hum Genet. 2018;103(4):535–52.

36. Köhler S, Bauer S, Horn D, Robinson PN. Walking the interactome for prioritization of candidate disease genes. Am J Hum Genet. 2008;82(4):949–58.

37. Cowen L, Ideker T, Raphael BJ, Sharan R. Network propagation: a universal amplifier of genetic associations. Nat Rev Genet. 2017;18(9):551–62.

38. Marees AT, de Kluiver H, Stringer S, Vorspan F, Curis E, Marie-Claire C, et al. A tutorial on conducting genome-wide association studies: Quality control and statistical analysis. Int J Methods Psychiatr Res. 2018;27(2):e1608.

39. Fadista J, Manning AK, Florez JC, Groop L. The (in)famous GWAS P-value threshold revisited and updated for low-frequency variants. Eur J Hum Genet. 2016;24(8):1202–5.

40. Machiela MJ, Chanock SJ. LDlink: a web-based application for exploring population-specific haplotype structure and linking correlated alleles of possible functional variants. Bioinformatics. 2015;31(21):3555–7.

41. Machiela MJ, Chanock SJ. LDlinkR: An R package for rapidly calculating linkage disequilibrium statistics in diverse populations. Frontiers in Genetics. 2018;9:157.

42. Zou Y, Carbonetto P, Wang G, Stephens M. Fine-mapping from summary data with the “Sum of Single Effects” model. PLoS Genet. 2022;18(7):e1010299.

43. Zondervan KT, Cardon LR. The complex interplay among factors that influence allelic association. Nat Rev Genet. 2004;5(2):89–100.

44. Boyle AP, Hong EL, Hariharan M, Cheng Y, Schaub MA, Kasowski M, et al. Annotation of functional variation in personal genomes using RegulomeDB. Genome Res. 2012;22(9):1790–7.

45. Vasylieva V, Arefiev I, Bourassa F, Trifiro FA, Brunet MA. Proteomics Can Rise to the Challenge of Pseudogenes’ Coding Nature. J Proteome Res. 2024;23(12):5233–49.

46. An Y, Furber KL, Ji S. Pseudogenes regulate parental gene expression via ceRNA network. J Cell Mol Med. 2017;21(1):185–92.

47. Qi Y, Wang X, Li W, Chen D, Meng H, An S. Pseudogenes in Cardiovascular Disease. Front Mol Biosci. 2020;7:622540.

48. Csárdi G, Nepusz T, Traag V, Horvát S, Zanini F, Noom D, et al. igraph: Network Analysis and Visualization in R. 2025.

49. Valdeolivas A, Tichit L, Navarro C, Perrin S, Odelin G, Levy N, et al. Random walk with restart on multiplex and heterogeneous biological networks. Bioinformatics. 2018;35(3):497–505.

50. Kanehisa M, Furumichi M, Sato Y, Kawashima M, Ishiguro-Watanabe M. KEGG for taxonomy-based analysis of pathways and genomes. Nucleic Acids Res. 2023;51(D1):D587–d92.

51. Yu G, Wang LG, Han Y, He QY. clusterProfiler: an R package for comparing biological themes among gene clusters. Omics. 2012;16(5):284–7.

52. Milacic M, Beavers D, Conley P, Gong C, Gillespie M, Griss J, et al. The Reactome Pathway Knowledgebase 2024. Nucleic Acids Research. 2023;52(D1):D672–D8.

53. Yu G, He Q-Y. ReactomePA: an R/Bioconductor package for reactome pathway analysis and visualization. Molecular BioSystems. 2016;12(2):477–9.

54. Yu G, Wang L-G, Yan G-R, He Q-Y. DOSE: an R/Bioconductor package for disease ontology semantic and enrichment analysis. Bioinformatics. 2015;31(4):608–9.

55. Schriml LM, Arze C, Nadendla S, Chang Y-WW, Mazaitis M, Felix V, et al. Disease Ontology: a backbone for disease semantic integration. Nucleic Acids Research. 2012;40(D1):D940–D6.

56. Jain A, Tuteja G. TissueEnrich: Tissue-specific gene enrichment analysis. Bioinformatics. 2018;35(11):1966–7.

57. Benjamini Y, Hochberg Y. Controlling the False Discovery Rate: A Practical and Powerful Approach to Multiple Testing. Journal of the Royal Statistical Society: Series B (Methodological). 1995;57(1):289–300.

58. Breunig MM, Kriegel H-P, Ng RT, Sander J. LOF: identifying density-based local outliers. SIGMOD Rec. 2000;29(2):93–104.

59. Collins RL, Glessner JT, Porcu E, Lepamets M, Brandon R, Lauricella C, et al. A cross-disorder dosage sensitivity map of the human genome. Cell. 2022;185(16):3041–55.e25.

60. European Organization For Nuclear R, OpenAire. Zenodo. CERN; 2013.

61. Quinodoz M, Royer-Bertrand B, Cisarova K, Di Gioia SA, Superti-Furga A, Rivolta C. DOMINO: Using Machine Learning to Predict Genes Associated with Dominant Disorders. Am J Hum Genet. 2017;101(4):623–9.

62. Hajdarevic R, Lande A, Mehlsen J, Rydland A, Sosa DD, Strand EB, et al. Genetic association study in myalgic encephalomyelitis/chronic fatigue syndrome (ME/CFS) identifies several potential risk loci. Brain Behav Immun. 2022;102:362–9.

63. Che X, Brydges CR, Yu Y, Price A, Joshi S, Roy A, et al. Metabolomic Evidence for Peroxisomal Dysfunction in Myalgic Encephalomyelitis/Chronic Fatigue Syndrome. International Journal of Molecular Sciences [Internet]. 2022; 23(14).

64. Huang K, G. C. de Sá A, Thomas N, Phair RD, Gooley PR, Ascher DB, et al. Discriminating Myalgic Encephalomyelitis/Chronic Fatigue Syndrome and comorbid conditions using metabolomics in UK Biobank. Communications Medicine. 2024;4(1):248.

65. Germain A, Barupal DK, Levine SM, Hanson MR. Comprehensive Circulatory Metabolomics in ME/CFS Reveals Disrupted Metabolism of Acyl Lipids and Steroids. Metabolites. 2020;10(1).

66. Hoel F, Hoel A, Pettersen IK, Rekeland IG, Risa K, Alme K, et al. A map of metabolic phenotypes in patients with myalgic encephalomyelitis/chronic fatigue syndrome. JCI Insight. 2021;6(16).

67. Xiong R, Gunter C, Fleming E, Vernon SD, Bateman L, Unutmaz D, et al. Multi-‘omics of gut microbiome-host interactions in short- and long-term myalgic encephalomyelitis/chronic fatigue syndrome patients. Cell Host & Microbe. 2023;31(2):273–87.e5.

68. Baraniuk JN. Cerebrospinal fluid metabolomics, lipidomics and serine pathway dysfunction in myalgic encephalomyelitis/chronic fatigue syndroome (ME/CFS). Scientific Reports. 2025;15(1):7381.

69. Quinville BM, Deschenes NM, Ryckman AE, Walia JS. A Comprehensive Review: Sphingolipid Metabolism and Implications of Disruption in Sphingolipid Homeostasis. Int J Mol Sci. 2021;22(11).

70. Bikman BT, Summers SA. Ceramides as modulators of cellular and whole-body metabolism. J Clin Invest. 2011;121(11):4222–30.

71. Sedlak TW, Saleh M, Higginson DS, Paul BD, Juluri KR, Snyder SH. Bilirubin and glutathione have complementary antioxidant and cytoprotective roles. Proc Natl Acad Sci U S A. 2009;106(13):5171–6.

72. Paul BD, Lemle MD, Komaroff AL, Snyder SH. Redox imbalance links COVID-19 and myalgic encephalomyelitis/chronic fatigue syndrome. Proc Natl Acad Sci U S A. 2021;118(34).

73. Chang CM, Warren JL, Engels EA. Chronic fatigue syndrome and subsequent risk of cancer among elderly US adults. Cancer. 2012;118(23):5929–36.

74. Francisco LFV, da Silva RN, Oliveira MA, Dos Santos Neto MF, Gonçalves IZ, Marques MMC, et al. Occupational Exposures and Risks of Non-Hodgkin Lymphoma: A Meta-Analysis. Cancers (Basel). 2023;15(9).

75. Bell DS. The Doctor’s Guide to Chronic Fatigue Syndrome: Understanding, Treating, and Living With CFIDS. Reading, MA: Addison-Wesley; 1995.

76. Costa DC, Tannock C, Brostoff J. Brainstem perfusion is impaired in chronic fatigue syndrome. QJM. 1995;88(11):767–73.

77. Miller AH, Jones JF, Drake DF, Tian H, Unger ER, Pagnoni G. Decreased basal ganglia activation in subjects with chronic fatigue syndrome: association with symptoms of fatigue. PLoS One. 2014;9(5):e98156.

78. Henrich MT, Oertel WH, Surmeier DJ, Geibl FF. Mitochondrial dysfunction in Parkinson’s disease – a key disease hallmark with therapeutic potential. Molecular Neurodegeneration. 2023;18(1):83.

79. Holden S, Maksoud R, Eaton-Fitch N, Cabanas H, Staines D, Marshall-Gradisnik S. A systematic review of mitochondrial abnormalities in myalgic encephalomyelitis/chronic fatigue syndrome/systemic exertion intolerance disease. J Transl Med. 2020;18(1):290.

80. Wang P-y, Ma J, Kim Y-C, Son AY, Syed AM, Liu C, et al. WASF3 disrupts mitochondrial respiration and may mediate exercise intolerance in myalgic encephalomyelitis/chronic fatigue syndrome. Proceedings of the National Academy of Sciences. 2023;120(34):e2302738120.

81. Evans DG, Harkness E, Woodward E. The TP53 c.455C>T p.(Pro152Leu) pathogenic variant is a lower risk allele with attenuated risks of breast cancer and sarcoma. Journal of Medical Genetics. 2023;60(11):1057–60.

82. Singh S, Kumar M, Kumar S, Sen S, Upadhyay P, Bhattacharjee S, et al. The cancer-associated, gain-of-function TP53 variant P152Lp53 activates multiple signaling pathways implicated in tumorigenesis. J Biol Chem. 2019;294(38):14081–95.

83. Morris G, Maes M. Increased nuclear factor-κB and loss of p53 are key mechanisms in Myalgic Encephalomyelitis/chronic fatigue syndrome (ME/CFS). Med Hypotheses. 2012;79(5):607–13.

84. Wang PY, Ma W, Park JY, Celi FS, Arena R, Choi JW, et al. Increased oxidative metabolism in the Li-Fraumeni syndrome. N Engl J Med. 2013;368(11):1027–32.

85. Perez C, Blomberg J, Gunter C, Cabanas H, Nacul L, Marshall-Gradisnik S, et al. Longitudinal cytokine and multi-modal health data of an extremely severe ME/CFS patient with hypermobility spectrum disorder. Frontiers in Immunology. 2024;15:1369295.

86. Carruthers BM, van de Sande MI, De Meirleir KL, Klimas NG, Broderick G, Mitchell T, et al. Myalgic encephalomyelitis: International Consensus Criteria. J Intern Med. 2011;270(4):327–38.

87. Gonzaga-Jauregui C, Lupski JR. Genomics of rare diseases. Walthum: Elsevier; 2021. pages cm p.

88. Gargano MA, Matentzoglu N, Coleman B, Addo-Lartey EB, Anagnostopoulos AV, Anderton J, et al. The Human Phenotype Ontology in 2024: phenotypes around the world. Nucleic Acids Res. 2024;52(D1):D1333–d46.

89. Thompson R, Papakonstantinou Ntalis A, Beltran S, Töpf A, de Paula Estephan E, Polavarapu K, et al. Increasing phenotypic annotation improves the diagnostic rate of exome sequencing in a rare neuromuscular disorder. Hum Mutat. 2019;40(10):1797–812.

90. Millard LAC, Davies NM, Gaunt TR, Davey Smith G, Tilling K. Software Application Profile: PHESANT: a tool for performing automated phenome scans in UK Biobank. Int J Epidemiol. 2018;47(1):29–35.

91. Uhlén M, Fagerberg L, Hallström BM, Lindskog C, Oksvold P, Mardinoglu A, et al. Tissue-based map of the human proteome. Science. 2015;347(6220):1260419.

92. Gómez-Mellado VE, Chang JC, Ho-Mok KS, Bernardino Morcillo C, Kersten RHJ, Oude Elferink RPJ, et al. ATP8B1 Deficiency Results in Elevated Mitochondrial Phosphatidylethanolamine Levels and Increased Mitochondrial Oxidative Phosphorylation in Human Hepatoma Cells. Int J Mol Sci. 2022;23(20).

93. Shi CH, Mao CY, Zhang SY, Yang J, Song B, Wu P, et al. CHCHD2 gene mutations in familial and sporadic Parkinson’s disease. Neurobiol Aging. 2016;38:217.e9-.e13.

94. De Rocco D, Cerqua C, Goffrini P, Russo G, Pastore A, Meloni F, et al. Mutations of cytochrome c identified in patients with thrombocytopenia THC4 affect both apoptosis and cellular bioenergetics. Biochimica et Biophysica Acta (BBA) - Molecular Basis of Disease. 2014;1842(2):269–74.

95. Harms FL, Girisha KM, Hardigan AA, Kortüm F, Shukla A, Alawi M, et al. Mutations in EBF3 Disturb Transcriptional Profiles and Cause Intellectual Disability, Ataxia, and Facial Dysmorphism. The American Journal of Human Genetics. 2017;100(1):117–27.

96. Cavdar Koc E, Burkhart W, Blackburn K, Moseley A, Spremulli LL. The Small Subunit of the Mammalian Mitochondrial Ribosome: IDENTIFICATION OF THE FULL COMPLEMENT OF RIBOSOMAL PROTEINS PRESENT *. Journal of Biological Chemistry. 2001;276(22):19363–74.

97. Lopez MB, Garcia MN, Grasso D, Bintz J, Molejon MI, Velez G, et al. Functional Characterization of Nupr1L, A Novel p53-Regulated Isoform of the High-Mobility Group (HMG)-Related Protumoral Protein Nupr1. J Cell Physiol. 2015;230(12):2936–50.

98. Camacho JA, Obie C, Biery B, Goodman BK, Hu C-A, Almashanu S, et al. Hyperornithinaemia-hyperammonaemia-homocitrullinuria syndrome is caused by mutations in a gene encoding a mitochondrial ornithine transporter. Nature Genetics. 1999;22(2):151–8.

99. Mayor A, Martinon F, De Smedt T, Pétrilli V, Tschopp J. A crucial function of SGT1 and HSP90 in inflammasome activity links mammalian and plant innate immune responses. Nat Immunol. 2007;8(5):497–503.

100. de Oliveira Otto MC, Lemaitre RN, Sun Q, King IB, Wu JHY, Manichaikul A, et al. Genome-wide association meta-analysis of circulating odd-numbered chain saturated fatty acids: Results from the CHARGE Consortium. PLoS One. 2018;13(5):e0196951.

101. Zhao B, Luo T, Li T, Li Y, Zhang J, Shan Y, et al. Genome-wide association analysis of 19,629 individuals identifies variants influencing regional brain volumes and refines their genetic co-architecture with cognitive and mental health traits. Nat Genet. 2019;51(11):1637–44.

102. Walter W, Sánchez-Cabo F, Ricote M. GOplot: an R package for visually combining expression data with functional analysis. 17 ed 2015. p. 2912–4.

103. Luo W, Brouwer C. Pathview: an R/Bioconductor package for pathway-based data integration and visualization. Bioinformatics. 2013;29(14):1830–1.

