## Supplementary Material for "Toward a Disease Module for ME/CFS: A Network-Based Gene Prioritization"

| Variant (h19) | rsid | beta | pval | PIP | RDB | Gene |
|---|---|---|---|---|---|---|
| 13:41353297:G:A | rs7337312 | -1.403E-03 | 2.56E-08 | 0.02 | 1f | - |
| 13:41358365:CT:C | rs368309529 | -1.514E-03 | 2.92E-09 | 0.14 | 2b | - |
| 13:41358369:T:C | rs564453195 | -1.514E-03 | 2.92E-09 | 0.14 | 2b | - |
| 13:41363595:T:C | rs7997189 | -1.391E-03 | 3.50E-08 | 0.02 | 1f | SLC25A15 |
| 13:41369790:A:G | rs1536807 | -1.449E-03 | 9.55E-09 | 0.05 | 1b | SLC25A15 |
| 13:41370996:A:G | rs6563846 | -1.456E-03 | 7.74E-09 | 0.06 | 1f | SLC25A15 |
| 13:41374328:C:G | rs2282023 | -1.457E-03 | 7.49E-09 | 0.06 | 1f | SLC25A15 |
| 13:41381077:G:A | rs2282026 | -1.456E-03 | 7.75E-09 | 0.06 | 1f | SLC25A15 |
| 13:41387065:T:A | rs2017696 | -1.492E-03 | 3.41E-09 | 0.12 | 1b | SLC25A15 |
| 13:41390881:G:A | rs7330720 | -1.485E-03 | 4.02E-09 | 0.10 | 1f | - |
| 13:41404706:T:C | rs11147812 | -1.530E-03 | 1.63E-09 | 0.23 | 7 | TPTE2P5 |

Table S 1. Statistically significant marginal regressions found between females with self-reported ME/CFS (phenotype 20002_1482) and common variants (minor allele frequency above 0.05). Beta is the coefficient of marginal logistic regression, and pval is the associated p-value. Regressions were performed by Neale Lab (24). PIP is the posterior inclusion probability assigned by *SusieR*. RDB is a rank assigned by RegulomeDB (version 2.2), and it indicates the probability of regulatory features (the lower the rank, the higher the probability).

| rs7337312 | rs368309529 | rs564453195 | rs7997189 | rs1536807 | rs6563846 | rs2282023 | rs2282026 | rs2017696 | rs7330720 | rs11147812 | Count | Frequency |
|---|---|---|---|---|---|---|---|---|---|---|---|---|
| G | T | T | T | A | A | C | G | T | G | T | 91 | 0.5 |
| A | - | C | C | G | G | G | A | A | A | C | 77 | 0.423077 |
| A | - | C | C | G | G | G | A | A | A | T | 6 | 0.032967 |
| G | T | T | T | G | G | G | A | A | A | C | 3 | 0.016484 |
| A | T | T | C | G | G | G | A | A | A | C | 2 | 0.010989 |
| A | T | T | C | G | G | G | A | A | A | T | 2 | 0.010989 |
| A | - | C | C | A | G | G | A | A | A | C | 1 | 0.005495 |

Table S 2. Frequencies of the haplotypes for the variants collected in Table S 1 in the GBR population. This table was calculated by function `LDhap()` of package *LDlink* (41) with hg19 as reference genome, GBR as reference population, and rsids of Table S 1 as input variants.

| | $L_1$ | $L_2$ | $L_3$ | $L_4$ | $L_5$ | $L_6$ | $L_7$ | $L_8$ | $L_9$ | $L_{10}$ | $L_{11}$ |
|---|---|---|---|---|---|---|---|---|---|---|---|
| $L_1$ | 1.0000 | 0.8877 | 0.8877 | 0.9157 | 0.8823 | 0.8958 | 0.8956 | 0.8961 | 0.8932 | 0.8938 | 0.7747 |
| $L_2$ | 0.8877 | 1.0000 | 0.9482 | 0.8918 | 0.8591 | 0.8731 | 0.8729 | 0.8734 | 0.8707 | 0.8713 | 0.7925 |
| $L_3$ | 0.8877 | 0.9482 | 1.0000 | 0.8918 | 0.8591 | 0.8731 | 0.8729 | 0.8734 | 0.8707 | 0.8713 | 0.7925 |
| $L_4$ | 0.9157 | 0.8918 | 0.8918 | 1.0000 | 0.8868 | 0.9002 | 0.9001 | 0.9006 | 0.8977 | 0.8983 | 0.7790 |
| $L_5$ | 0.8823 | 0.8591 | 0.8591 | 0.8868 | 1.0000 | 0.9326 | 0.9324 | 0.9329 | 0.9301 | 0.9307 | 0.8120 |
| $L_6$ | 0.8958 | 0.8731 | 0.8731 | 0.9002 | 0.9326 | 1.0000 | 0.9458 | 0.9463 | 0.9434 | 0.9440 | 0.8257 |
| $L_7$ | 0.8956 | 0.8729 | 0.8729 | 0.9001 | 0.9324 | 0.9458 | 1.0000 | 0.9461 | 0.9432 | 0.9438 | 0.8256 |
| $L_8$ | 0.8961 | 0.8734 | 0.8734 | 0.9006 | 0.9329 | 0.9463 | 0.9461 | 1.0000 | 0.9437 | 0.9443 | 0.8260 |
| $L_9$ | 0.8932 | 0.8707 | 0.8707 | 0.8977 | 0.9301 | 0.9434 | 0.9432 | 0.9437 | 1.0000 | 0.9414 | 0.8233 |
| $L_{10}$ | 0.8938 | 0.8713 | 0.8713 | 0.8983 | 0.9307 | 0.9440 | 0.9438 | 0.9443 | 0.9414 | 1.0000 | 0.8239 |
| $L_{11}$ | 0.7747 | 0.7925 | 0.7925 | 0.7790 | 0.8120 | 0.8257 | 0.8256 | 0.8260 | 0.8233 | 0.8239 | 1.0000 |

Table S 3. Linkage disequilibrium matrix for the variants of Table S 1. Element *i,j* of the matrix was calculated as $f_{i,j} - f_i f_j$ divided by the square root of $f_i f_j (1 - f_i)(1 - f_j)$, where $f_i, f_j$ are the frequencies of the alternative alleles of the control group at positions $i, j$ respectively, and $f_{i,j}$ is the frequency of the haplotype defined by the presence of the alternative alleles at positions $i$ and $j$, in the reference population (42). The frequencies of the haplotypes for this table are derived from Table S 2.

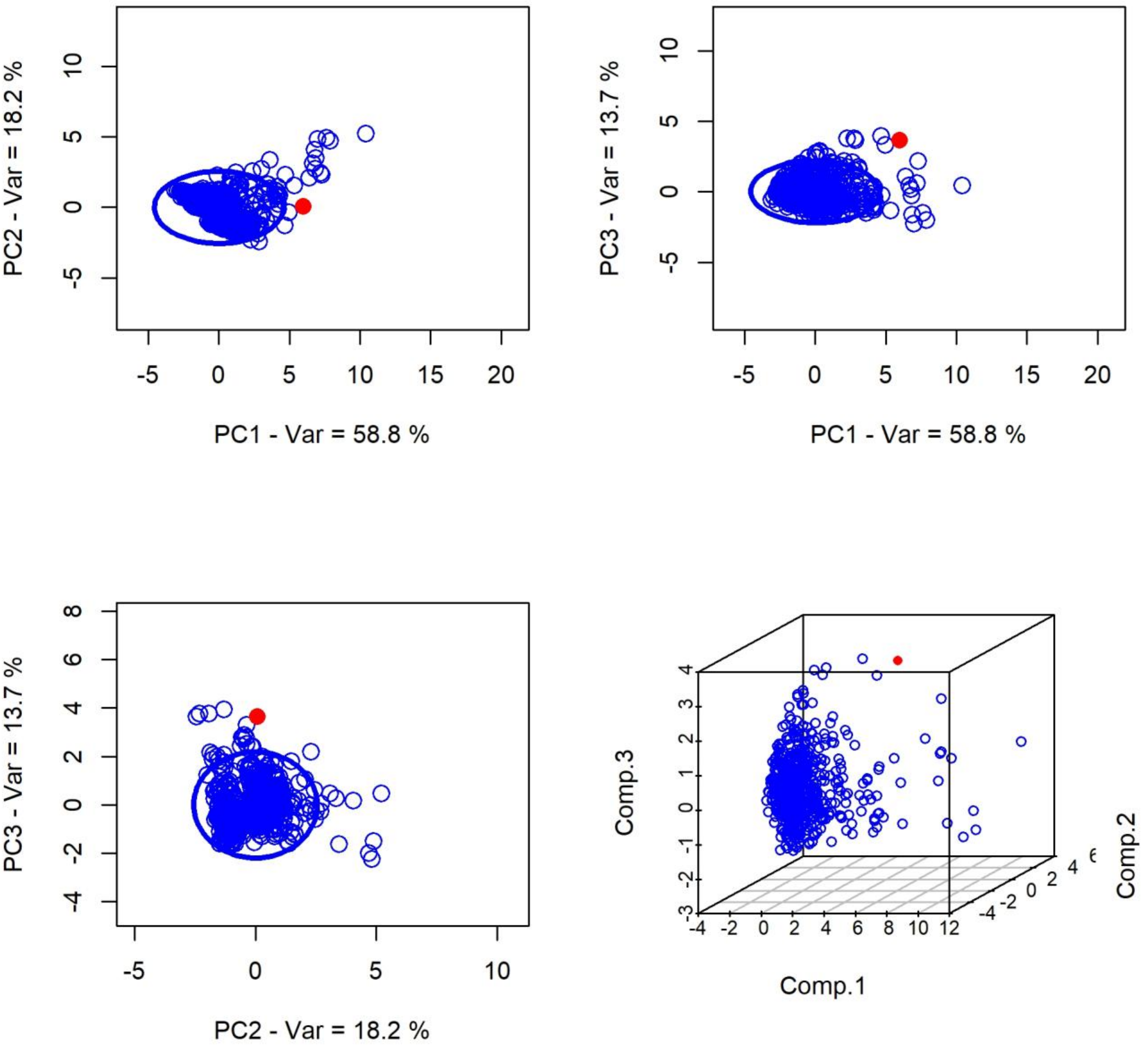


Figure S 1. Principal component analysis (PCA). Plane PC1-PC2 (top-left), plane PC1-PC3 (top-right), plane PC2-PC3 (bottom-left), and space PC1-PC2-PC3 (bottom-right). The dot representing the merged gene list (MGL) associated with ME/CFS is indicated in red, while the blue dots refer to six hundred MGLs built from random seed genes. The variables considered are as follows. GG_components: inverse of the number of connected components of the gene graph; GG_nodes: number of nodes of the gene graph; LG_components: inverse of the number of connected components of the list graph; LG_edges: number of edges of the list graph; LG_weight: total weight of the edges of the list graph; LG_degree: mean degree of the nodes of the list graph. The red dot falls outside the 95% ellipse in each one of the three principal planes considered here. This figure was generated by a custom R script.

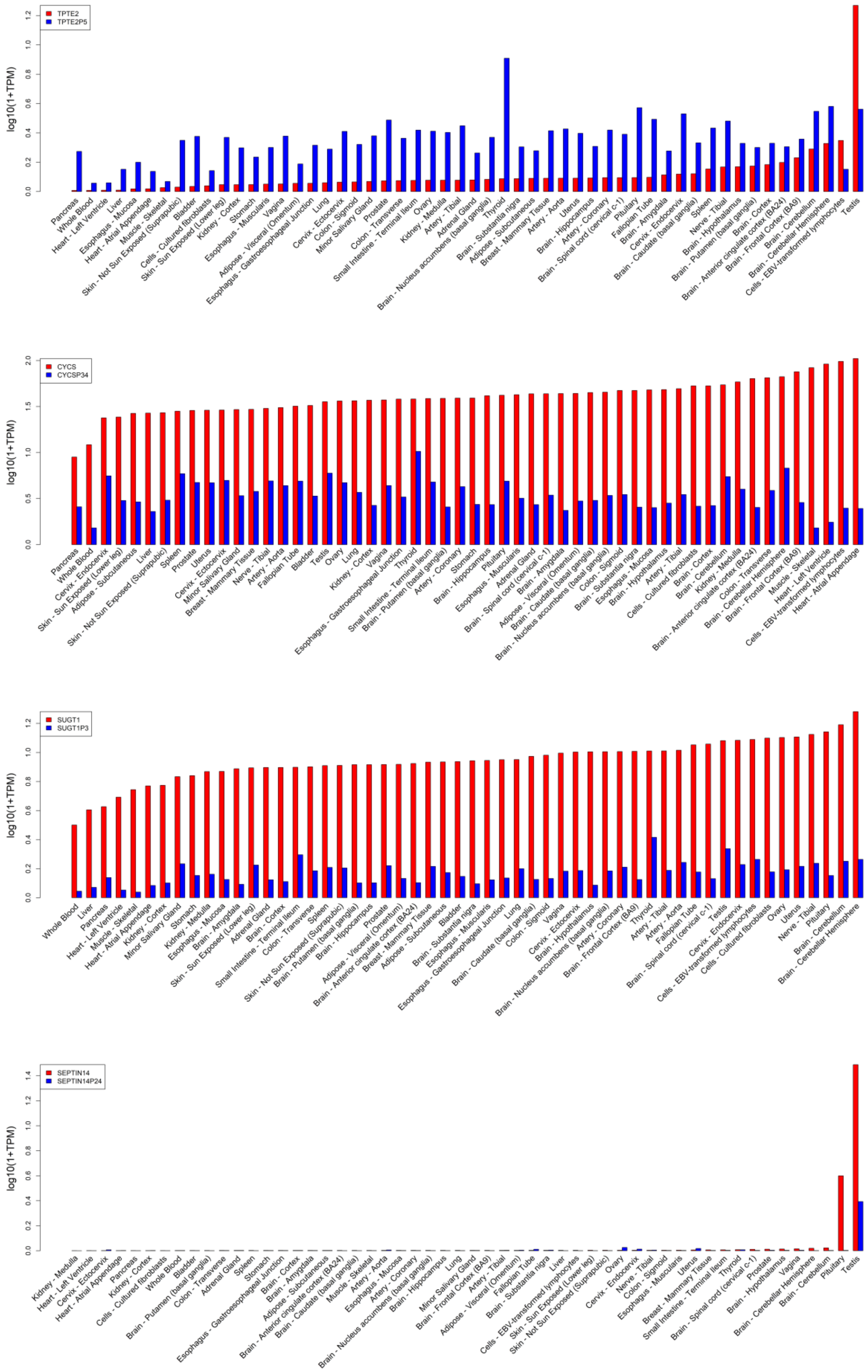


Figure S 2. Gene expression by tissue of pseudogene-parental gene pairs, reported as $\log_{10}(1 + \mathrm{TPM})$. Note that this is not absolute gene expression; therefore, the expression of one gene/pseudogene cannot be directly compared with the expression of another.

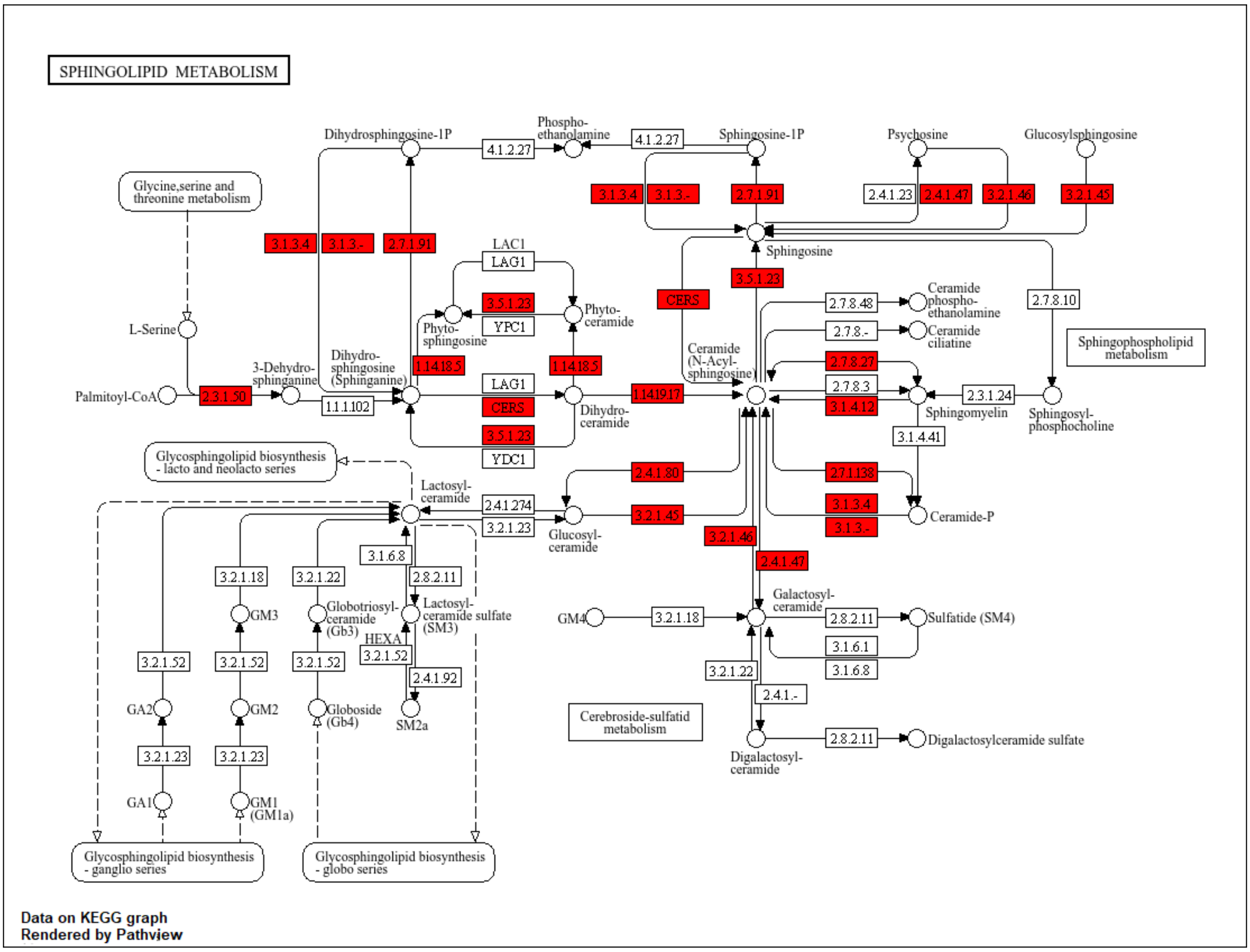


Figure S 3. Sphingolipid metabolism. This KEGG pathway diagram highlights in red the genes from the ME/CFS module that map to the sphingolipid metabolism pathway (hsa00600). The pathway illustrates the enzymatic steps involved in the synthesis and degradation of sphingolipids, including ceramide and sphingomyelin. The enrichment of this pathway in the ME/CFS gene module supports previously reported abnormalities in lipid metabolism, particularly of ceramides and sphingomyelins. Visualization was generated using the package *Pathview* (103). The thirty-two genes of the ME/CFS module that overlap with this pathway are collected in Supplementary Table S4 and Figure 7.

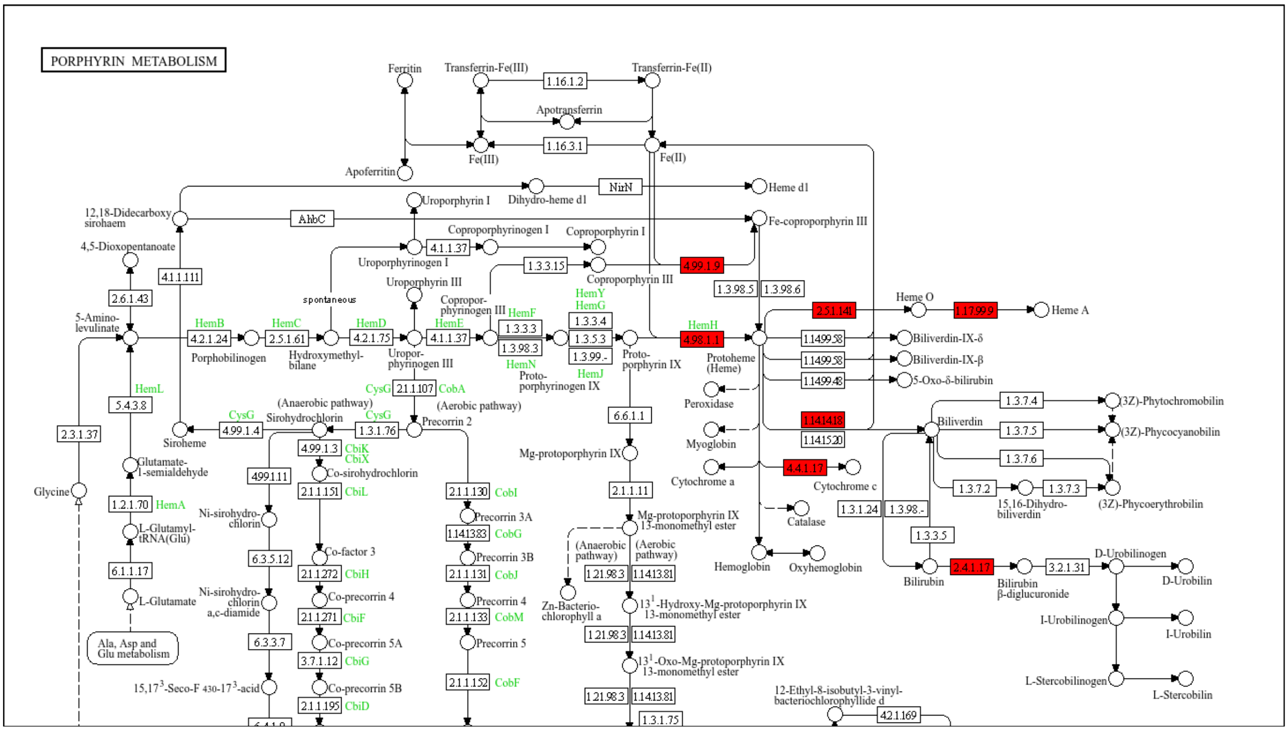


Figure S 4. Porphyrin metabolism. This KEGG pathway diagram highlights in red the genes from the ME/CFS module that overlap with the porphyrin metabolism pathway (hsa00860), specifically clustering in the heme degradation branch. These genes participate in the enzymatic conversion of heme into biliverdin, bilirubin, and other metabolites. This aligns with clinical observations of reduced bilirubin levels and increased heme in patient cohorts. Visualization was generated using the package *Pathview* (103). The original pathway diagram was cut in the lower half to show only the part that overlaps with the ME/CFS module. The twenty-three genes of the ME/CFS module that overlap with this pathway are collected in Supplementary Table S4 and Figure 7.

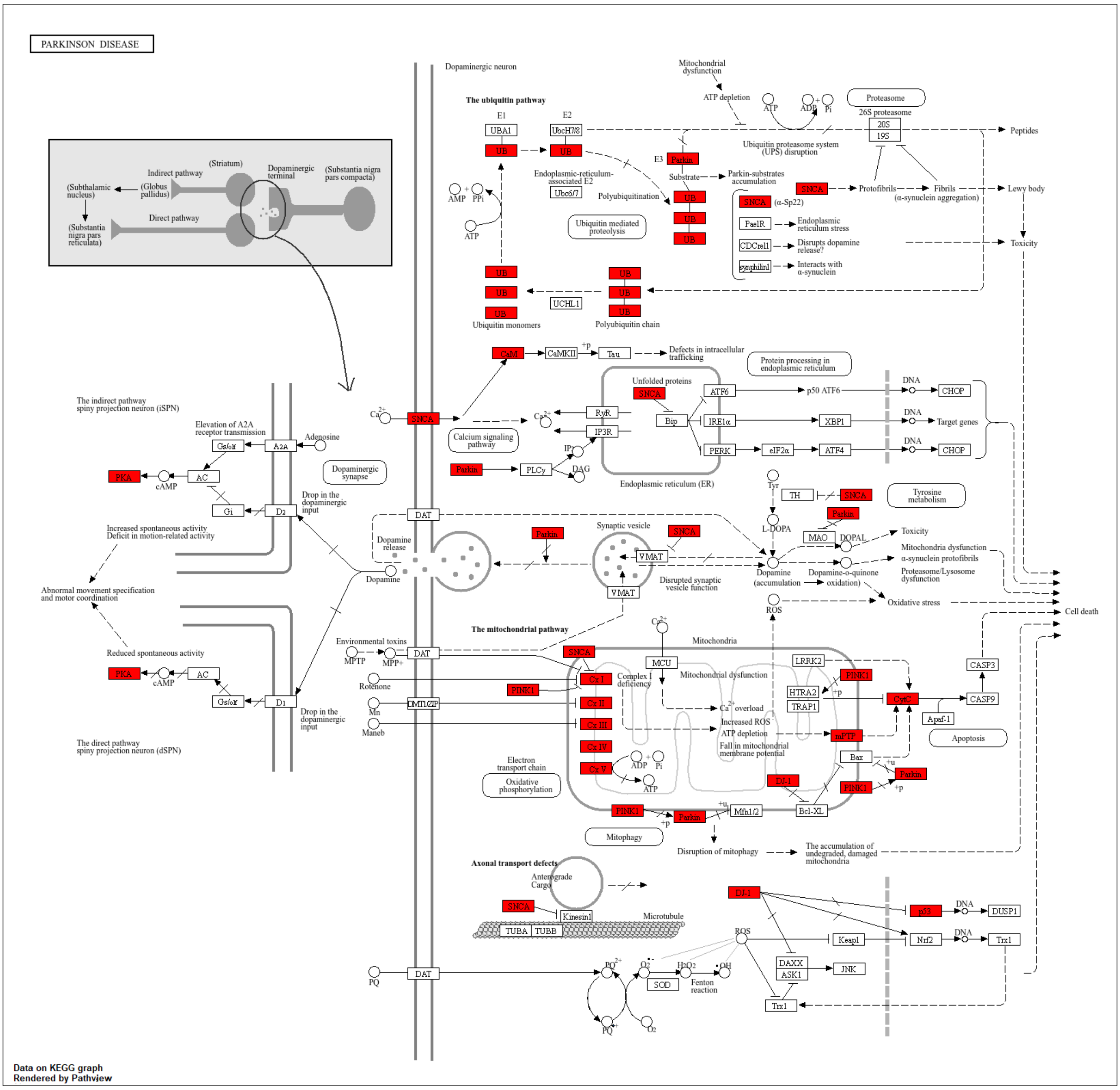


Figure S 5. Parkinson's disease. This KEGG pathway diagram highlights in red the ME/CFS module genes that overlap with the Parkinson's disease pathway (hsa05012). These genes participate in oxidative phosphorylation, proteasomal degradation, and dopaminergic synapse regulation. Visualization was generated using the package *Pathview* (103). The forty-four genes of the ME/CFS module that overlap with this pathway are collected in Supplementary Table S4 and Figure 7.

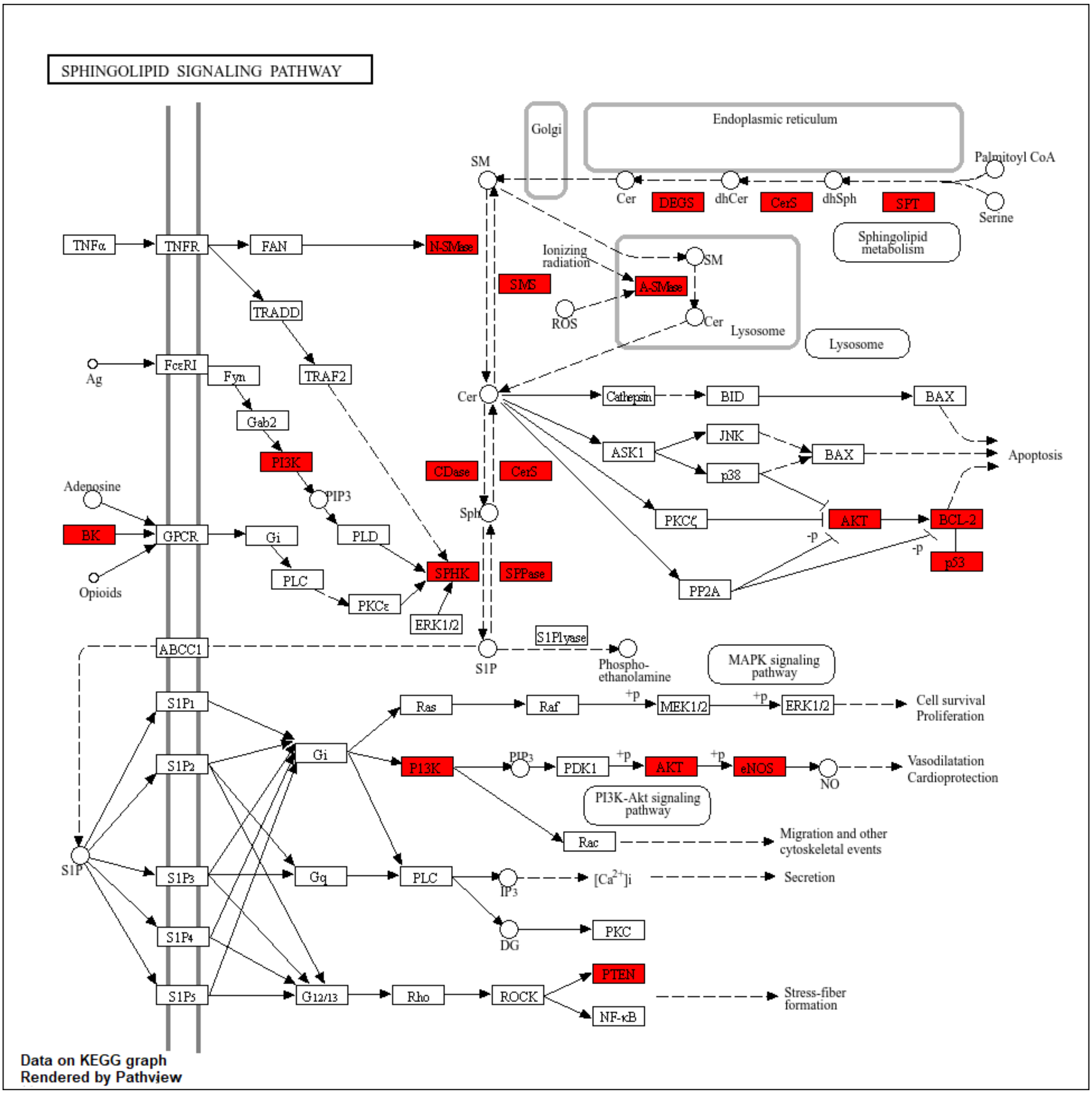


Figure S 6. Sphingolipid signaling. This KEGG diagram highlights in red the ME/CFS module genes that overlap with the sphingolipid signaling pathway (hsa04071). These genes participate in the synthesis and downstream signaling of ceramide, sphingosine (Sph), and sphingosine-1-phosphate (S1P), which regulate diverse cellular processes, including apoptosis, inflammation, stress response, cell survival, and mitochondrial function. Visualization was generated using the package *Pathview* (103). The thirty genes of the ME/CFS module that overlap with this pathway are collected in Supplementary Table S4.

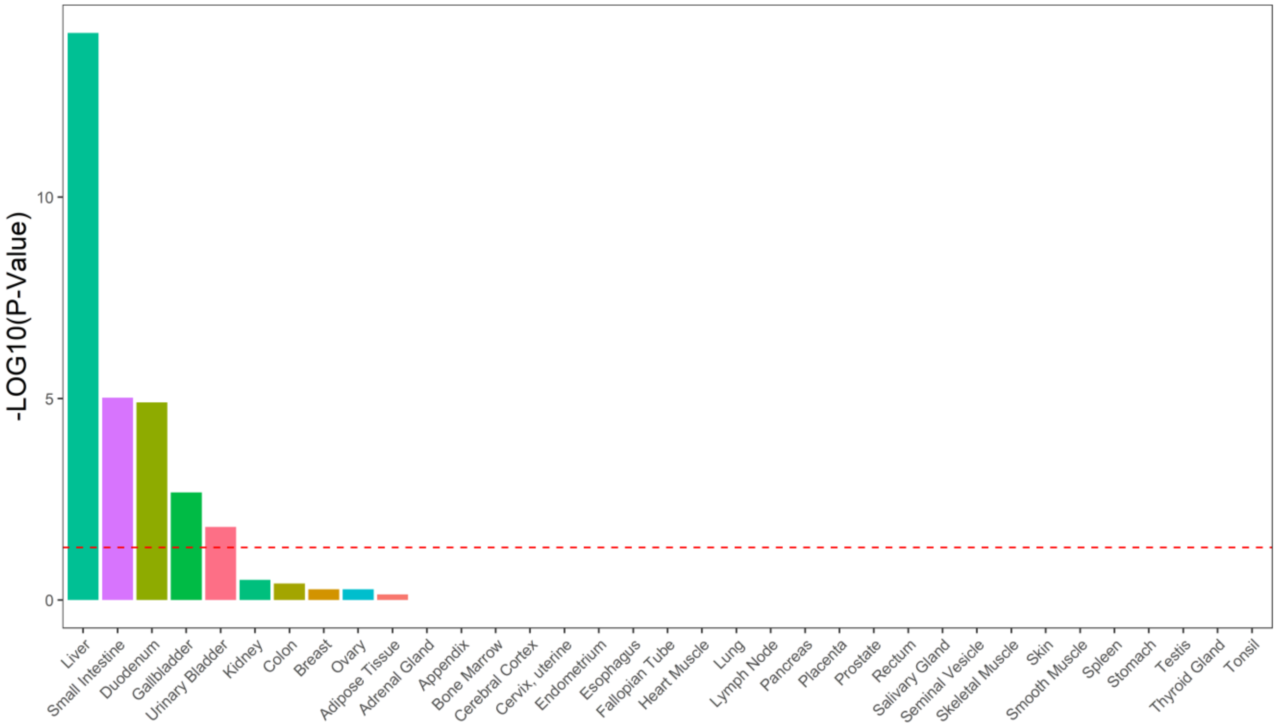


Figure S 7. Tissue enrichment of the ME/CFS module. Bar plot showing the $-\log_{10} p$ of the enrichment of tissue-specific genes in our ME/CFS module. Enrichment was performed on the Human Protein Atlas database by function `teEnrichment()` of package *TissueEnrich* (56) with a correction for multiple comparisons (Benjamini-Hochberg method). The horizontal dashed line indicates the cut-off for significance: $-\log_{10} 0.05 \sim 1.3$.
